# Tracking the tension: Examining emotional conflict experienced in wearable activity tracker users

**DOI:** 10.64898/2025.12.03.25341327

**Authors:** Gabrielle Humphreys, Sam Jensen, Ashley Gluchowski

## Abstract

Wearable activity trackers have been recognised as effective tools for physical activity promotion, leading to their integration in healthcare services. Although, some qualitative literature indicated that device users may experience emotional conflict. The current study is the first of our knowledge to directly examine the conflict faced by wearable activity tracker users.

A qualitative, exploratory design was followed, with inductive thematic analysis conducted on semi-structured interview transcripts. The current study consisted of 11 participants (8 female), aged between 18 to 59 years (M= 30.73) who used a wearable activity tracker for a minimum of 3 months.

Four themes and nine sub-themes captured participants’ emotional conflict. Themes were; Who knows best? Who’s in charge? Who am I without it? And What is happening to me?. Themes highlighted that device users faced emotional conflict around navigating a data mismatch, how a wearable activity tracker impacted their behaviour, the amount of control a tracker had over them, whether their device use was positive, and how they would act and feel if they no longer used their wearable activity tracker Participants experienced substantial emotional conflict from wearable activity tracker use. The intensity of device-user relationship was clear, suggesting device dependency and perceived device importance. Findings hold crucial implications around the integration of activity trackers in healthcare services, recommendations around healthy use, and the potential long-term negative impact of using these devices on bodily intuition. Theoretical underpinnings remain unclear around wearable activity tracker use; results suggested blurred boundaries between intrinsic and extrinsic motivation - likely due to device embodiment - and highlighted the role of pressure in driving increased physical activity.

**Author summary:** Wearable activity trackers allow users to self-track health information. Typically devices are watches, but rings and chest straps are also used. We investigated if individuals using these devices experienced any emotional conflict. Interviews were conducted and data was analysed. We found that users felt emotional conflict for multiple reasons. Many received data from their device that they disagreed with, meaning they had to debate which perspective to adopt. Others felt conflict around how important their device felt to them and how much it impacted their behaviour. These devices were worn consistently by most participants meaning they became dependent on a device and found it difficult to separate what behaviour was for themselves, and what was to please the device. Consistent use meant users felt conflict around who they would feel and act if they stopped using their activity tracker. Conflict was also reported around how these devices changed behaviour; some said a device increased their motivation, but many instead reported feeling pressure to be active. Wearable activity trackers are beginning to be used in healthcare services. These findings stress that we need to recommend them with caution.

## 1. Introduction

### 1.1 Context

Currently standing as one of the fastest growing sectors in the technology industry [1], 37% of UK individuals surveyed currently own and use a wearable activity tracker (WAT) [2], with this most popular in 35 to 44 year olds [3].

Worldwide, there are an estimated 454 million smartwatch users – a subtype of WATs – with rates increasing yearly [4].

Wearable activity trackers (WATs) are portable electronic monitoring devices which allow individuals to self-track their daily physical activity [5]. Self-tracking often involves continuous monitoring of in-depth bodily information, with the idea of users gaining control to optimise their health behaviours, resulting in the concept of a quantified self [6]. Henriksen et al. [7] identified 423 devices with Fitbit, Garmin, Misfit, Apple and Polar as leading brands.

These devices measure and feedback data on step count, distance moved, heart rate, level of activity, and sleep patterns [8], with features falling under the functional domains of monitoring and nudging [9]. WATs typically embed gamification into their user experience, often featuring badges, streaks, and leaderboards to promote both physical activity and WAT engagement [10].

Some WATs include smartphone-based features, coined smartwatches, whereas others allow self-tracking without messaging services.

### 1.2 WATs as behaviour change interventions

While these devices were first perceived as lifestyle accessories, wearable activity trackers are now viewed as a legitimate tool for health promotion and ill-health prevention by healthcare professionals [11, 9]. The majority of quantitative research considers WATs through the lens of interventions, examining their effects on objective measures such as physical activity [12]. In their umbrella review of 39 papers across 163,992 participants, Ferguson et al. [12] examined the impact of WATs on physical activity and health outcomes in all age ranges and both clinical and non-clinical groups. Here, the use of WATs was significantly associated with improved physical activity, body composition, and fitness. Specifically, using these devices resulted in an additional 1,800 steps, and 40 minutes of walking per day, resulting in participants’ body weight reduced by approximately 1kg. Notably, quantitative data around behavioural outcomes tends to be mixed with overall results often averaging out as neutral responses [13].

When examining mechanisms behind this health promotion behaviour change, motivation has been discussed heavily in literature [14–18]. Jung and Kang [19] suggested these WATs specifically increase intrinsic motivation of users, highlighting the application of the self-determination theory (SDT) within this context. Within SDT, fitness data tracking was positively related to user autonomy, and in turn, feelings of enjoyment [19]. Organismic Integration Theory, a sub-theory within SDT focussing on the extrinsic motivation types [20] has been used to explain how WATs lead to physical activity promotion in users [21]. Specifically, James, Wallace and Deane [21] reported different preferences in WAT features between intrinsically and extrinsically motivated users; intrinsically motivated preferred social features, whereas extrinsically motivated preferred exercise control features (e.g., exercise prompts, goal management). Although, the COM-B model [22] was also identified as a plausible explanation to WAT use [23, 24], alongside 28 other theoretical frameworks, suggesting current lack of clarity around this behaviour [24]. This lack of clarity may exist given the plethora of individuals’ relationships with health and reasons for engaging in WAT use, which in turn may lead to different behaviour change experiences [21].

### 1.3 The emotional impact of WATs

Qualitative studies in this area have gained traction aiming to understand the multifaceted concept of digitalised health. In their meta-synthesis of 18 studies Sandham et al. [25] concluded that receiving data on health improvement since using a WAT increased user motivation to continue this positive trajectory.

Nelson et al. [26] reported that motivation levels peaked three-months following device acquisition due to novelty being replaced with device familiarity, whereas Burford, Golaszewski and Bartholomew [27] concluded that motivation was increased long-term, driven by continuous feelings of accountability and feelings of intrinsic motivation from WAT feedback.

Although, in some cases participants appeared to rely on this information for validation, resulting in device embodiment [26]. Nelson et al. [26] compared participants’ WAT-body integration similar to a prosthetic limb, highlighting how natural using these devices became for users, and how dependent some were on them. Similarly, Köhler et al. [9] shared that participants felt this was no longer a removable accessory; suggesting device embodiment and dependence.

However, contrasting user experiences were common amongst these qualitative studies. Burford, Golaszewki and Bartholomew [27] highlighted that feedback led to distress in some users. Specifically, one user reported that this feedback *‘made or broke their day.’* Another participant took their smartwatch off on the weekends when step count was lower to avoid feedback, reporting that they *‘can’t handle it’*; highlighting the perceived importance of this feedback. Device frustration was also experienced by users, specifically over receiving nudges to move during inflexible periods [17]. In their systematic review of females’ experiences with WATs, Del Busso et al. [18] highlighted the pressure experienced by some individuals to be active and continuously improving.

While some studies in this review reported overwhelmingly positive findings, in 3 of the 13 studies reviewed, women reported that perceived stress and pressure from WATs resulted in device avoidance [28–30].

Individual conflict around WAT use has appeared in previous literature, although not directly examined. For example, in Toner, Allen-Collinson and Jones’ work, a participant noted the ‘mental trade off’ they perform when receiving a prompt to move (Stan, 35) [17]; navigating conflict around pros and cons of movement. This same participant reported their WAT has led to them doubting their own ability to judge the quality of their sleep, with feedback leading to data mismatches, and in turn, emotional conflict.

Perceived fairness, accuracy, and value of feedback determined user reactions to this information, with some users facing what has been coined a data mismatch or data-expectation gap [31,32,25]. This occurs when a mismatch between device detection and expected output has occurred, leading to feelings of frustration and mistrust when experienced [31]. Frustration over forgetting to track exercise or a devices battery running out was reported; further emphasising the importance users placed in obtaining their data, and thus, the conflict faced if a data mismatch was experienced.

### 1.4 The current study

This study is the first paper directly examining emotional conflict of individuals using wearable activity trackers.

Building upon the conflict mixed emotional feelings reported within Toner, Allen-Collinson and Jones [17], this study poses the research question, how do individuals experience and navigate emotional conflict when using smartwatches? With WATs being integrated into healthcare settings to encourage behaviour change [32–34] and initial guidance on WAT usage emerging [35], this work holds important implications.

To gain this insight, the following objectives were proposed;

1. Explore the emotional experiences of smartwatch users in relation to activity tracking, performance feedback and daily monitoring.
2. Examine the emotional conflict experienced by smartwatch users in regard to receiving this health feedback.
3. Understand the emotional and behavioural impact of this emotional conflict.
4. Understand how individuals overcome this emotional conflict.

## 2. Methods

### 2.1 Study design

The current study holds a qualitative, exploratory design using semi-structured interviews to examine this topic. A reflective, inductive thematic analysis approach was adopted [36]. This allowed for an in-depth understanding into this complex topic of psychological experiences to data provision, building on our current knowledge on wearable health trackers.

### 2.2 Participants (selection and recruitment)

Participants were aged 18+ individuals who reported at least three-months of consistent wear of an activity tracker. Consistent use was defined as wearing the device almost all of the time, allowing for charging periods. Three months was selected as a minimum duration to ensure that participants had established routines and familiarity with device feedback, allowing reflection on both novelty and emotional impact.

While smartwatches are the most popular type of wearable activity trackers (defined by their integration of messaging and calling features) [37], all WATs were examined in the current study. Examples of Garmin, Coros, Fitbit, Apple, Samsung and Google Pixel watches were listed, alongside Whoop bands and Oura rings. No restrictions were placed upon the brand of wearable trackers, the amount of physical activity completed, or other demographics to promote a heterogenous sample.

The study was advertised in multiple ways. Physical posters were displayed around a northern university where the research team were based. Digital posters were displayed via platforms relating to digital health – specifically via the university portal, LinkedIn and X.

Recruitment continued until data saturation was reached, with 11 participants in the final sample.

### 2.3 Data collection

After providing informed consent for the current study, interviews were arranged. All interviews were conducted by GH, either in-person (n= 4) or online via Microsoft Teams (n= 7). Interview durations ranged from 19 to 82 minutes (M= 48). Interviews were semi-structured, with prompts used to encourage conversation. Questions first asked about demographic data and an overview of WAT use. Discussion then focussed on the impact of smartwatch data on behaviour and emotions, prompting for any conflicting experiences. Next, participants were asked about their perceived accuracy and importance of smartwatch data, prompting for any experienced conflict. Following interview questions, participants received a verbal debrief, reminded of researcher contact details and their right to withdraw. All interviews were recorded via Microsoft Teams and automatically saved to the primary researchers’ password protected OneDrive account.

### 2.4 Data analysis

Interviews were transcribed automatically through Microsoft Teams, and then quality checked by researchers. Braun and Clarke’s [36] six stages of thematic analysis were used to analyse data. Data was first coded independently by two researchers (GH; SJ; stages 1 to 3) to prevent biases. Coding was completed using Microsoft Word to annotate text. A reflexive, inductive approach was adopted ensure all content was encapsulated from the dataset. Coding was iterative, with the research team revisiting transcripts multiple times as understanding developed. GH and SJ then reviewed their codes; defining and naming themes collaboratively. AG and KM reviewed interview transcripts and the table of quotes to ensure proposed themes well represented conversation. All identified quotes were written into a table (see Supporting Information 1), with those most relevant discussed in the main body of this paper. A COREQ checklist was used to ensure transparent reporting of this study [37].

### 2.5 Reflexivity and research team

The positionality of the research team was considered throughout this project, aiming for any personal biases around this topic to be minimised. The team consisted of four academics across multiple disciplines (psychology, sport, and nursing) to ensure wide perspectives. GH and AG used a WAT consistently, KM used a WAT selectively for exercise only, and SJ had never owned a WAT.

No qualities about the research team, other than the university affiliation of the research project, were reported to participants. Although, the study was advertised across the university meaning some may have been aware of a professional background (n=3 undergraduate students).

### 2.6 Ethical statement

Ethical consideration such as anonymity, potential relationships between participants and researcher, and sensitive discussion topics were considered. Ethical approval was gained from the University of Salford ethics board in June 2025; reference 6343. Written informed consent was obtained from all participants prior to interviews being conducted.

## 3. Results

### 3.1 Participant characteristics

11 participants were recruited for the current study. The sample consisted of eight females and three males, a mean age of 30.73 years (SD = 11.65), and included a range of ethnicities with White-British most common. Participants’ current WATs spread across five brands, with Garmin the most often used. See Table 1 for participant characteristic breakdown. Identifiable information was removed from any discussed quotes and pseudonyms were used in write up.

**Table 1.** Participant characteristics.

| Participant | Age | Gender | Ethnicity | Device |
| --- | --- | --- | --- | --- |
| 1 | 31-34 | Female | White – Australian | Garmin Forerunner 265 |
| 2 | 26-30 | Female | White – British | Apple Watch Series 8 |
| 3 | 18-25 | Female | White – Spanish | Garmin Forerunner 265 |
| 4 | 26-30 | Male | White – British | Garmin Fenix 6 X Pro |
| 5 | 35-40 | Male | Asian - British | Garmin Fenix 5 X |
| 6 | 55-60 | Female | White – British | Fitbit Charge 4 |
| 7 | 18-25 | Female | White – British | Apple Watch Ultra 2 |
| 8 | 18-25 | Female | White – British | Apple Watch Series 6 |
| 9 | 35-40 | Female | White - Swedish | Garmin Fenix 7 Pro |
| 10 | 35-40 | Male | Black - British | Coros Pace Pro |
| 11 | 18-25 | Female | White – British | Apple Watch (model unknown) |

### 3.2 Thematic analysis

Four themes were identified to reflect participants’ experiences of emotional conflict in WAT use; **Who knows best?**, **Who’s in charge?**, **Who am I without it?** and **What’s happening to me?**. Within these were nine sub-themes (see Figure 1).

**Figure 1.**
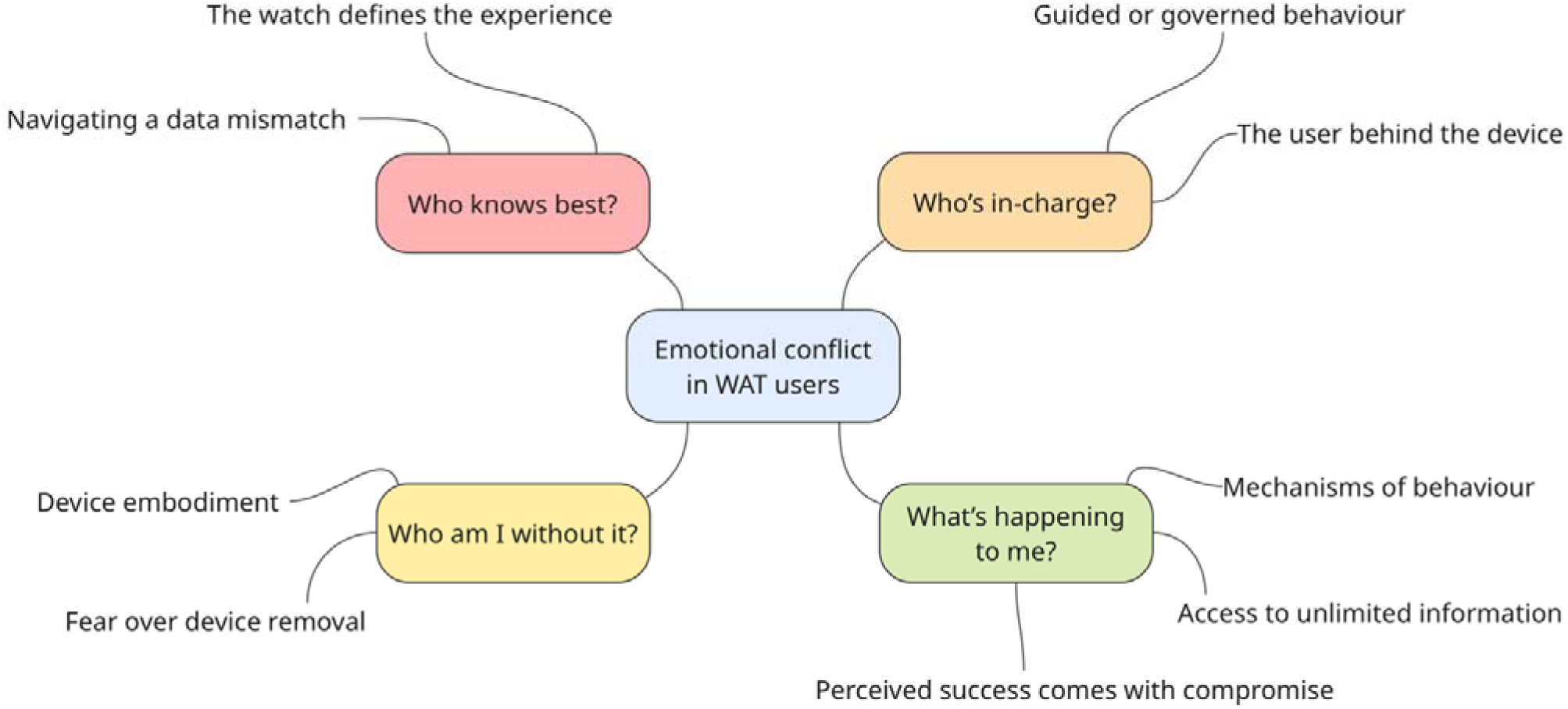
Thematic map.

#### Theme 1. Who knows best?

Who knows best? highlighted the perceived expertise of WATs, with the user seeking their device to define the experience over their own intuition. If data provided by a WAT was not aligned with users’ perception of an activity, this required users to navigate a data mismatch; either believing their own perceptions or adopting the digital viewpoint.

#### The WAT defines the activity

A WAT defined an activity via the data it provided. Data provided prior to performing an activity predicted, and potentially determined, the experience; *‘I was looking at training readiness and training status to tell me like almost how I’m gonna do at the gym’ (ZA),* and also applied to receiving feedback following an activity. Without this data, users thought exercising would *‘feel like it hadn’t counted’ (MT)*. This belief was despite participants acknowledging no physical change had occurred, for example, they *‘still walked their dog and literally did the same routes’ (JP)*, yet they *‘wouldn’t want to workout without a watch on’ (MT)*, therefore, they would feel *‘pretty angry if it died during a workout’* (AH) Participants reported valuing data over their own experience, specifically feeling more informed; *‘I obviously would know like I went in at this time and came out at this time, but I wouldn’t know how hard I pushed’ (MT)*. This meant WAT users *‘see people working out without tech and [are] like but what do you mean, like, like how do you know what you did?’ (MN)* therefore relying on digitalised feedback over intuitive bodily feedback. This resulted in participants embedding data into conversation; *‘The first thing me and my partner always do is ask “how did you sleep” and we both then look at our sleep on the watches and talk about it’ (SA). Participant AH also reported this behaviour; ‘I go how did you sleep and he’ll say “yeah, 92” and then I’ll be like oh, what! I only got 81”’*.

Some participants relied on data as their goals were numbers-based, such as taking 10,000 steps per day, in which they *‘could have been on 8,000 without a watch’*, therefore, *‘they wouldn’t know if I’d met my targets without the data’ (MT),* or running a certain distance*;‘I could take it off but how would I know that I ran 5k?’ (MN).* In this case, the device provides *‘a lot of statistics that [they] basically need in order to see if [they’re] improving or not’ (SA)*.

Although, other participants wanted data for a broader level of recognition; *‘If I didn’t use it I would have no idea what kind of activity I was completing’ (HG)* highlighting WAT data was viewed as necessary to define an activity over feeling. This was echoed frequently by participants; *‘I don’t know how I’d know what I’d done in a day’ (MT); ‘how would I know what I did?’ (ZA)*; so much so, participants had been mocked about their data reliance; *‘I said to my auntie, I was like I feel naked, I don’t have my watch on, and she was like how will you know if you’re alive?!*’ *(MN)* demonstrating the visible impact of WATs to a non-user.

Notably, users placed considerable trust in a WAT, *‘feel[ing] like it knows that better than me’ (MH)* and had such a drive for objectivity in their performance, they questioned the purpose of activity without this data; *‘if I don’t have statistics…like what am I doing it for?’ (SA)*. Similarly, if data was negative, participants thought ‘what’s the point?’ *(MH)*, suggesting that WAT users overlooked the value of physical activity for health and well-being, instead viewing the benefit as a positive digital score. With this mindset, performing an activity without an activity tracker *‘doesn’t feel the same, even though I know it absolutely is’* therefore, participants thought *‘well, I can’t go anywhere now or do like any steps because they don’t count’ (HG)*. Participant JP acknowledged this *‘sounds absolutely ridiculous’*, yet, this was their experience of digitalised versus non-digitalised movement.

Although, this stance was not consistently adopted both between and within participants. While Participant JP stated importance around tracking their step count, they questioned *‘I don’t know what I’d do with the fact I recorded a swim’ (JP).* Participant MH adopted a similar stance around stress-related data, saying *‘I don’t need a watch to tell me how stressed I am’ (MH).* Furthermore, Participant LP planned to change their WAT use, sharing *‘I’m listening to it too much. I can just get out and do it and then I will judge how it was based on my experience instead of what a watch says’ (LP).* Instead, they currently *‘use [the data] to judge how [they’ve done]…seeing it as this score that comes above everything else’*, whereas *‘without that number [they’d] feel good’ (LP)*.

#### Navigating a data mismatch

If users disagreed with WAT feedback they had to navigate a data mismatch; ultimately deciding whether to adopt the viewpoint of a WAT or themselves. Participants reported experiencing data mismatches around sleep; *‘I might feel well rested and then I’ll see I got a 62 out of 100’ (AH)*, and heart rate; *‘it shows my heart rate is really low…I think I’ve done a good workout and it’s just not showing that (ZA).* Mismatches also arose following activity tracking, for example, when *‘you do like a workout or like a walk or something, it gives you an effort level, but it’s not tailored, you don’t have any say’ (HG)*, with a WAT reporting the user as *‘strained when [they] feel strong or the other way round’ (ZA)*.

Given the trust placed in WATs, when navigating a data mismatch users typically adopted the device’s perspective. For example, Participant MT said *‘There’s going to be a reason behind the numbers it’s pumping out and you get a watch to learn more…I’m not going to argue with it and tell it it’s wrong when it’s a tool, like, it’s tracking me’ (MT).* Participant MH echoed this, saying ‘*as a person you’re only capable of considering so many variables at one time, whereas [a WAT has] got the full picture and it’s privy to, considering variables that you’re not necessarily even aware of’*. This adoption of the WAT’s viewpoint often occurred due to the perceived accuracy in tracking compared to the individual alone, for example, an individual *‘can’t count to 28 minutes’ or ‘can’t measure 5k’*, therefore meaning *‘you’ve got to go with [the WAT] there’ (ZA)*.

In some mismatches participants shared they ‘debate it out’ (LP), comparing this investigation to *‘if your average uni grade was like 70 and then you got 52…you would question it’ (MT).* In these cases being hungover, tired, ill, or the weather being hot were examples of context users applied to try explain their data output. Although, other participants were quicker to dismiss their own perceptions. For example, Participant AH *‘had such a deep sleep’* but receiving a sleep score lower than expected, stated *‘I never know if then I see that score and it unconsciously makes me feel more or less tired’ (AH),* concluding she *‘takes [the WAT] on board [and] listen to it more than [herself] which is pretty terrible’.* Similarly, LP concluded *‘oh well, I must have actually slept badly’ (LP)*, highlighting the adoption of data as one’s own feelings. Participant MH added they *‘would always lean to trusting the watch’* because it is *‘so much more concrete than feeling*’. This suggests that for many, facing a data mismatch simply means your perceived experience was incorrect.

In contrast, others reported rejecting the WAT’s perspective when navigating a mismatch. Participant HG stated *‘I know my body better than that’*, expanding that a WAT *‘just doesn’t sync to like life events very well…expecting every day is going to be the same’ (HG).* Others aimed to *‘take what it tells me with a pinch of salt’ (MH)* and be *‘guided by what my body is doing rather than what the watch is telling me’ (MN)*. Although, Participant MN noted that adopting this mindset was not easy, with data still making them *‘second guess’* and *‘absolutely question’* their experience. Importantly, adopting this mindset appeared to be a learned behaviour developed over time, specifically formed after *‘taking it too far in the other direction’ (LP)*. This was needed *‘because when you start trying to exercise more and stuff, you rely on it because you don’t know any better’ (HG)*. This balanced mindset was *‘something that [MN] had to develop over time and not have the tech rule my life…maybe that’s an age thing’ (MN)*.

Importantly, a data mismatch sometimes resulted in a positive outcome with WATs providing *‘rational information [AH] needed when [their] brain has been devilish and on fire and really quite harsh, it’s given [them] that objective information’ (AH).* Similarly, if the WAT feedback was more positive than expected, JP reflected *‘That can’t be true, but I just get excited’* because *‘If you don’t feel very fit and it tells you you’re amazing…oh, what a lovely compliment! (AH).* Inputting data into a WAT created a digital diary, highlighting to KL *‘I haven’t actually felt that awful all week. I’ve been OK. It’s just right now I don’t feel good’.* Participant AH shared a similar narrative; *‘I go on a run and I double take at the speed I’m doing. I almost don’t believe it and I know for a fact without that watch I wouldn’t feel as strong and healthy, then empowered to do more’ (AH).* This highlighted that in some cases, a data mismatch could provide emotional reassurance rather than correction, showing that trust in data can both soothe and unsettle. However, negative outcomes typically arose from navigating a data mismatch. Participants would *‘rip [their] hair out’ (MH)* becoming *‘furious from the lack of agreement’ (AH),* even finding a mismatch *‘kind of offensive’ (HG).* Participant AH reflected on discovering their watch provided inaccurate data around running pace. This left them questioning their ability and sent them *‘into a real spiral’*, asking *‘what if I think I’m alright at this and it turns out all that data was wrong? I’m actually not fast at all?’*; and transitioning to running on a treadmill over a three-month period; *‘so I could see the actual data’.* While some participants felt low levels of confusion and stress, this highlights the huge impact WAT reliance can have on a user from both an emotional and behavioural perspective.

#### Theme 2. Who’s in-charge?

While who knows best? explored conflicts of knowledge, who’s in-charge? considers how WATs shape a users’ behaviour. Participants consistently acknowledged that their wearable activity trackers shaped everyday behaviour, yet uncertainty surrounded who directed this change. This theme proposed two alternative perspectives; firstly, that the watch guides or governs behaviour, or, that the user behind the device is responsible for the impact.

#### Guides or governs behaviour

Reflecting on how a WAT impacted behaviour – through guiding or governing – came with difficulty. This relationship lacked clarity with Participant AH saying *‘I try pick out the reasons why I want to do something. Is it for me or is it for the watch? Sometimes I’m not sure’ (AH).* Some participants used the watch to inform small, healthy behaviours which contributed to a larger lifestyle; *if I’d not done much I could, say, park far away from the shops’ (JP)* or after noticing a low step coun*t ‘walk to the shops instead of driving’ (MH).* Sleep data also informed energy levels with Participant SA *‘scheduling a cycle race earlier rather than later’* if they slept poorly, and previous activity informed future activity; *‘I didn’t get my move goal so this week…I’ll use my walking pad when I’m working’ (HG)*.

In contrast, others said *‘the watch makes decisions for me’ (JP)*. Participant KL said *‘I’m getting controlled by a watch is basically what I’m telling you’,* adding *‘it’s like a little tyrant on my wrist and I’m forgetting that I’ve got free will’ (KL)*. Participant LP shared they have *‘debated stopping wearing one because [they] find it’s just like [they’re] listening to it a bit too much’*, so much so, they felt like they’d *‘almost lost [they’re] own autonomy’(LP).* The perceived expertise of these devices meant participants would use these devices to provide permissions of behaviour and set parameters for their lives; ZA shared *‘using it for essentially knowing what I’m doing each day’.* Participant MT highlighted the extent this data can govern behaviour, saying; *‘I just use it all the time. I use it for my eating…I use it to plan should I go for a walk or should I sit on the sofa, or yeah, like what meal to eat that night, I look at the calories I’ve got left’ (MT).* Participant ZA highlighted the permission a device provided around calorie consumption also, where data *‘will let me know what I can have’ and ‘decide what I can do’ (ZA).* Notably, they acknowledge that this relationship *‘doesn’t sound healthy at all’* and shared they *‘don’t like saying I look at my watch to know what to eat’ (ZA).* Users found their WAT *‘might sometimes make me exercise or do stuff when I don’t actually feel like it or when my body doesn’t really want to’ (MN),* such as *‘doing steps around [their] kitchen…trying to walk about more to get steps up’ (ZA).* Participant LP questioned this behaviour, sharing *‘I think the thing I’m struggling with is that you get a smartwatch to change your behaviour…but now…I’m questioning like if a watch should be changing my behaviour. It definitely changes my behaviour, I just don’t know how I feel about it’ (LP)*, highlighting that while this adaptation to a WAT governing behaviour is desired for some, it can be a concern for others. This impact extended beyond WAT users too, as ZA and his partner had a *‘massive lie in’*, resulting in ZA repeatedly *‘checking the time because it was like 2:00pm and we’d not done anything and my step count was basically zero, so I was thinking yeah I probably need to’ (ZA).* This resulted in ZA’s partner questioning his behaviour and calling the device *‘toxic’*.

#### The user behind the WAT

While a WAT seemed to change behaviour, the impact of the user on this relationship must be questioned. Participants were goal focussed; *‘I set myself lots of goals anyway. Love a goal. I’ve always been quite goal driven’ (LP)* and committed to their health; keeping a *‘wellness journal’ (SA)* of activities before owning a WAT. In this case, a WAT didn’t introduce a new mindset around exercise, instead simply changing manual tracking to digital;*‘instead of you keeping track of what you’re doing there’s a device that does it for you’ (SA)*. Although, given the data a WAT provides, *‘the watch means goals are based on numbers’ (LP).* Therefore, while goals may have been more intuitive and subjective in non-WAT users, WAT users seemed to be *‘anal and single focussed’ (MH)* on this data. Participants saw their performance as a *‘score’ (ZA)* they wished *‘to beat’ (MN),* reporting they *‘really like to see the numbers tally up’ (AH)*, suggesting a WAT gamified their health. Given participants were *‘pretty competitive’ (MT)*, when they did achieve this, participants felt *‘really excited’ (MN)*, although for some, simply *‘the chasing of the feeling [a new record] is enough’ (MN)*.

Some users had a negative relationship with exercise prior to WAT ownership, with this device then providing data to further emphasise effects; for example, Participant AH fixated on the calories burned during exercise as a *‘weird count up’*, and remarked *‘I know it was actually my fault as a user’ (AH)*. Similarly, participant MT said *‘I don’t know if that’s the watch or me with like exercise as a whole’ (MT)* over their attitudes to exercise. The impact of the WAT versus the user was difficult to decipher with users questioning; *‘but then like, is that the watch doing that or is that just my personality? Like can be blame the watch?’ (MN)*. Participant ZA added *‘it’s hard to know if I’ve put that pressure on myself or the watch has’*. Participant MH further emphasised the difficulty in separating the two, suggesting the device and user are two interacting components which cannot distinctly be separated; *‘It’s hard though cos I think that pressure is like internal, but it’s pressure from the knowing of the watch. I think what I’m trying to say is that someone else could get that data and do nothing with it’*.

#### Theme 3. Who am I without it?

Who am I without it? captures how participants described their wearable devices as extensions of themselves. Many participants reported device embodiment, where the device wasn’t just worn but incorporated into identity. For some, this relationship produces a sense of dependency where the removal of the device produced feelings of fear and anxiety.

#### Device embodiment

Participants frequently described their watches as inseparable from their sense of self: *‘it’s so close I feel like I can’t separate it from myself’ (HG)* and sharing that *‘It’s weird…it’s really difficult to separate between what is me and what is the watch’ (MH).* Participant AH supported this, asking *‘is it for me or is it for the watch? Sometimes I’m not sure’ (AH).* This suggests a form of device embodiment, where technology becomes incorporate into awareness and identity. Two participants in this sample had Type 1 Diabetes using monitors that were *‘integrated into the watch’ (HG)*. This context somewhat justified device embodiment. Yet, other participants, did not face these additional health needs but reported device embodiment, stating they were *‘not sure where it ends’ (LP)* when trying to separate themselves from their device.

Given their repeated use of these devices *‘for so many years’ (LP)* and having *‘never stopped using a smartwatch since [they] got it’ (AH)*, imagining not using a WAT anymore *‘would be weird’ (AH)*. Participants said they *‘feel naked’* if their device needs charging because *‘it just goes everywhere’ (KL)* with them. Constant device use was *‘habitual’ (CF)*, with Participant CF sharing *‘I’m so used to having it. It can be dead and I’ll still go to check it’ (CF)*, meaning if a WAT was removed this habit would *‘take a lot of reversing’ (LP)*. When discussing device removal Participant LP responded *‘it might be boring [and] a bit lonely, because I really like waking up and seeing what I’ve done. It’s like a paper in bed to me and it’s every day’.* This questioned whether users formed a parasocial relationship with their WAT from it adding a social element to their day. Participant SA highlighted the strength and intensity of this formed relationship, stating *‘I think I’d be actually lost without it’,* indicating that data no longer just represented activity, instead becoming part of how users understood themselves.

#### Fear over device removal

Given their device embodiment, participants expressed fear over device removal. Participant SA *‘would cry’* because *‘I wouldn’t have a reference point for if I was doing well’*; suggesting fear arose because WAT defined user experience and a form of emotional reassurance. Similarly, Participant MT was asked about exercising without their WAT and replied *‘I just wouldn’t…like even now you can tell I’m just so stressed’,‘I said absolutely not, there was no question about it…I wouldn’t workout without wearing one…It makes me feel sick’ (MT).* This highlighted the device dependency some participants, using emotive language and appearing visibly stressed when asked about this hypothetical scenario. Others shared *‘I’d probably panic if I didn’t have it’* stated Participant AH, despite knowing *‘I’m still doing the same thing even if it’s on or off my wrist’.* MN explained ‘*it comes back to that thing my auntie said, like, how will you know if you’re alive, how will you know your heart is beating?;* suggesting the loss of body intuition from a WAT. They also faced conflict over removal, acknowledging when exploring this use, adding the data *‘does make me anxious sometimes. It literally makes me anxious but the idea of not wearing it also freaks me out’ (MN),* indicating that participants recognised this dependency but felt powerless to resist it. These reactions could be explained due because constant data had become the norm; *‘When you’re given so much data, then the idea of having that data taken away just makes you feel like how do I know what I’m doing?’ (HG)*. Having feedback became *‘kind of obsessive’* which Participant HG *‘didn’t realise…until you said about taking it off. I was like I don’t want to do that!’.* This reflected the dependency many faced without realising due to their device embodiment.

#### Theme 4. What is happening to me?

What is happening to me? examined the effects of using a WAT. This considered the mechanisms behind behaviour with differing responses to data reported, alongside the impact of having access to unlimited information with a granular level of detail. Participants also justified experiencing negative effects from their WAT, explaining their perceived success comes with compromise. Positive data reinforced effort and pleasure, while negative scores provoked guilt, anxiety, or self-criticism.

#### Mechanisms of behaviour

Differing mechanisms of behaviour were reported by participants. Some behaviour change occurred by WATs increasing motivation;*‘you get such a boost because you feel like you’re doing well, then you invest more time into it’ (SA)*. Similarly, MH said their WAT created a *‘feedback loop’*, describe themselves as *‘a little rat who gets more cocaine at the end of a workout’* explaining *‘I get this endorphins kick after exercise, but then…I can look at the numbers and get an extra pat on the back’*. This highlighted the drive for positive feedback following a workout which, in turn, prompted motivation to repeat this behaviour, because *‘‘If it tells me I’ve done well, I feel good, stating the obvious right?’ (LP)*. Encouragement was also provided when close to reaching a goal also, for example, saying *‘you’ve made like three quarters of the progress, you’ve only got this bit left and that is really motivating’ (HG).* Then when a goal was met, participants felt *‘a sense of achievement’ (JP)* with evidence to say *‘wow, look at my steps!’*.

Anticipated positive feedback also motivated users. Participant KL shared thinking *‘if I go to sleep now my Apple Watch will tell me I’m doing good in the morning.’*. If they slept well, KL reported feeling *‘great because I know I’m getting a good score in the morning’.* Although, motivation was not the only mechanism involved here, with KL facing *‘the opposite effects’* if they can’t sleep, knowing ‘*it’s going to tell me off in the morning’ (KL)*. KL added *‘if I’ve forgotten to put my watch on for the night and then I’m struggling to sleep, it’s almost kind of a relief because…I’m not going to get told off in the morning’*; highlighting feeling pressure to perform well. Additionally, if having a late night, they *‘will purposefully take [their WAT] off just so [their] graph still looks nice’ (KL)*, suggesting that gaining positive feedback may be more of a priority than the behaviour itself. Participant SA also actively avoided negative feedback; when showering they *‘tend to remove it just because [they’re] worried my average heart rate on the day will go up’*.

Feelings of pressure or stress resulted in behaviour change for some; *‘I panic when sometimes I look at my training status and it says something like maintaining’* shared Participant SA, with this prompting them to exercise. ZA felt conflicted as *‘I really enjoy looking at the data but can sometimes find myself panicked on numbers…when nothing has actually changed in the last year, I just have the information now’ (ZA);* therefore, behaviour change may occur through perceived pressure to achieve continuous health improvement, with this improvement defined by WAT data alone.

Users reported the outcome of WAT feedback *‘depends on [their] mood’ (MH)*, saying *‘if I’m doing well it can be really motivating, but if you’re not, well, it can just be the total opposite’ (HG)*. AH similarly said *‘it depends on my mindset and where my head is cause if I see I’ve done something like 200 steps, or I’ve not worked out in a week…that’s when it could be guilt’*; suggesting low mood was only worsened by data. Although, feedback content seemed to be a larger determiner of response, with ZA stating *‘it basically depends on what the feedback is’*. This meant the mechanisms behind behaviour *‘depends on how you’re doing, you know, if it’s an active day or not. If it is, you feel great and it’s an extra pat on the back, but if you’re having to have a slow say it feels like a reminder that I’m not doing great.’ (JP).* This was echoed by Participant MT who said *‘it makes me feel really good…when I’ve met my goals, but if I’ve not met my goals yet, it makes me feel really stressed because it’s just this, like, flashing deadline’ (MT).* Therefore, the WAT was a *‘double-edged sword’ (JP)* amplifying the behaviour the user has done; either bringing further reward or pressure. *‘On one side it says get up lazy, but then the data on the other side says look at what you did’* supported AH. With feedback from a WAT therefore determined by the users behaviour, Participant SA hypothesised that *‘if you have a chronic condition, it might not be the best, but if you are healthy it can do so much good’*.

#### Perceived success comes with compromise

Participants acknowledged negative impact from WAT use, but explained their perceived success comes with compromise. For example, data made Participant ZA *‘quite anxious*’, although he described it as*‘a mean teacher that I don’t like, but then I’ve opened my results and think you know what, they knew what they were doing’ (ZA)*, justifying these negative experiences overall. The extent of this negative impact was notable with Participant MN struggling to think of any positive emotions; *‘I don’t think there are any like positives from day-to-day’ (MN)*, although ‘*while there could be all of these negative impacts, wearing a smartwatch makes [them] healthier’ (AH)*, suggesting users saw this emotional trade off as reasonable in order to meet behavioural goals. Participants perhaps overlooked these consequences because they bought a WAT to improve fitness; *‘it can be a little unhealthy, but the watch is having the effect I wanted’ (ZA)*.

This brought conflict with users registering the dislike for the device, yet enjoyment over the outcomes; resulting in thoughts that were *‘messy to unpack’ (JP)*. AH also questioned this, because *‘on paper it might have made me physically healthier’,* but it equally made them *‘obsessive with numbers’ (AH)*. Overall, users seemed to prioritise their behavioural goals over well-being, with Participant ZA acknowledged that not using their WAT would make them *‘a bit more relaxed [because] it wouldn’t tell me off’*, but concluded that is *‘not what I need right now’* because they *‘wouldn’t be as fit’ (ZA)*.

#### Having access to unlimited data

Owning a WAT means users have access to unlimited information. This was viewed as potentially harmful with Participant JP stating *‘Too much information isn’t good in my eyes’,* explaining *‘I keep it simple and I think that’s why I’m okay’.* Others did not limited their data consumption and reported *‘I think I just know a bit too much from it’ (ZA),* sharing *‘I’m obsessed with my heart rate, probably to the detriment of my anxiety’ (MN)*. Constant data may prevent users from switching off, with Participant AH stating *‘body freedom and just not having to think about things every single day is something I can’t really imagine’ (AH)*. Although, given the constant data provision from a WAT, body awareness made sense. Additionally, the granular level of detail only added to stress, with participant ZA comparing it to *‘doomscrolling and fearmongering’*. This detail also *‘makes you forget about the bigger picture. It doesn’t matter that I slept 6:50 and I’m really happy about that. Instead it kind of makes you zone in on the small things’* said KL.

Importantly, participants highlighted the long-term impact of this. Participant MH said *‘I sometimes wish I didn’t know’* things, but *‘the cats out the bag, I can’t unsee that now’ (MH)*. They added *‘ignorance is bliss’*, whereas their WAT made them *‘so aware of the impact [alcohol] has on my sleep and overall training readiness’* - knowledge they cannot reverse.

Given this long-term impact, participant ZA thought ‘*getting rid of my watch smartwatch would mean I’d just be trying to estimate [the data themselves] and then panicking about my guesses’ (ZA)*. ZA further elaborated saying ‘*now I’ve worn one I know the impact of movement. I just don’t know if I’ve done 5,000 or 6,000 steps so I’d only start guessing I reckon…I’d probably be more tired by thinking like that’ (ZA)*. Participant LP also described the long-term mindset that using a WAT created. Debating whether to remove their WAT, they shared *‘I feel like the watch is like the devil on my shoulder, or wrist’* because *‘my brain is going but what, but what about if you miss a PB?!’ (LP)*. Given users have gotten used to a WAT defining their experience, it appeared future activities without a device would result in disappointment that there was no onscreen confirmation of this activity.

A table of all relevant quotes is accessed in the Supporting Information 1.

## 4. Discussion

### 4.1 Key findings and existing literature

The current study built on the concept of the quantified self [6] which arose in WAT users, and answered James et al.’s [21] call to explore data mismatches.

Exploring this emotional conflict, four themes and nine sub-themes were identified; Who knows best?, Who’s in charge?, Who am I without it? And What’s happening to me?. Data depicted that intense relationships had formed between the WAT and user, in which the user valued WAT feedback highly and used it to make decisions. Given the frequency of use and trust placed in devices, some users expressed a loss of autonomy, bodily intuition and fear around life without their WAT.

Mechanisms of behaviour were examined within the sub-theme What’s happening to me?. WATs increased motivation to lead an active lifestyle in some participants, strengthening the conclusions of existing literature [14–18,25]. The current findings depart from previous literature by examining the user experience in depth. Uncertainty was expressed around how a WAT led to behaviour change, specifically around the type of motivation experienced.

Positive WAT feedback motivated users to continue their health-related behaviour; strengthening the findings of Burford, Golaszewski and Bartholomew [27] who reported that feeling validated via positive WAT feedback increased intrinsic motivation. This aspect of our findings supports the COM-B model, with feedback highlighting improvement likely to increase reflective motivation, and in turn, fuel the target behaviour [22]. Although, participants also detailed their WAT increased activity via feeling pressured which may not directly fit this model of behaviour. One could reframe feeling pressure to receive positive WAT feedback into being motivated to avoid negative feedback to map findings onto the COM-B model. However, this does not fully capture the authority these devices had over user’s behaviour and the drive participants expressed over avoiding negative feedback rather than performing the behaviours such as sleep and physical activity for their benefits. This drive to avoid negative feedback led to participants hacking their feedback by removing devices at times, with these users reporting satisfaction over this despite not meeting their personal goals.

These experiences highlighted a WAT consisted of conflicting intervention functions simultaneously; incentivisation was provided if users gained positive feedback, but coercion was experienced with many users performing behaviours simply to avoid being *‘told off’*. These findings are notable, supporting the conclusions of an umbrella review Ferguson et al. [12] that WATs increase physical activity, whilst stressing the importance of examining the mechanisms behind this behaviour change. All participants in the current study were full-time WAT users, and none had intentionally paused their device use. The theme perceived success comes with compromise showed that users felt a negative emotional impact was justified for improved health. However, current WAT users were only recruited in this study. Valuable future insight could be gained from those who no longer use a WAT, particularly given the reporting that stress from WATs can lead to device avoidance [28–30].

Participants expressed difficult understanding their behaviour given device embodiment had occurred; strengthening this concept proposed by Nelson et al. [26]. Nelson et al. [26] reported device use becoming natural behaviour and leading to dependence; findings matched in our study. Similarly, Köhler et al. [9] reported that participants no longer viewed WATs as removable accessories.

This was encapsulated within the theme Who am I without it?, with sub-themes of device embodiment and fear over device removal.

Device embodiment perhaps explained why mechanisms of behaviour were difficult for users to unpick, suggesting a blurring of intrinsic and extrinsic motivation. Furthermore, WATs may target different forms of motivation simultaneously. In their study on current wearable fitness trackers, Nuss and Li [39] found that users scored highly on both introjected (avoiding punishment, feelings of guilt) and integrated (congruence with one’s values, feelings of volition) forms of motivation. The present study corroborates these findings, as some participants reflected on the pressure to meet the goals outlined by their WAT, but other perceived such goals to align with internal goals. Furthermore, a prominent finding in the present study was how initial motivations behind the purchase of a WAT was that it aligned with participants’ pre-existing exercise habits or values around the importance of exercise. However, with time, this more autonomous form of motivation was influenced by feelings of pressure to meet goals. This was highlighted by Nuss and Li [39] amongst former users, who note that WATs can result in decrease in autonomous motivation due to external rewards and feedback, engaging with the WAT out of pressures to complete goals rather than personal choice. Similarly, Steel [40] found that WAT users also transitioned from more autonomous forms of motivation to introjected. Previous research identified the importance of WATs in quantifying participants physical activity [39], resulting in decreased motivation to exercise when tracking is not available [41]. Current findings support this, with participants reporting physical activity as *‘not counting’* if it has not been recorded; a concept labelled as the dependency effect [27].

However, in contrast to previous research that identifies the ways WATs support autonomy [19,37], many participants in the presented study reflected on how their needs for autonomy were blocked by their WAT. Jung and Kang [19] posit that a user’s control over the WAT and how data is used supports feelings of autonomy. Yet, whilst WATs have been discussed in their ability to provide users with choice over goals or activity type, the present study indicates that WATs are perceived to govern participants’ behaviour, to the point where a loss of their own autonomy was reported.

Participants often trusted device data over bodily intuition, so much so they questioned how they’d know anything without their WAT data. This highlighted a major consequence of the quantified self, with users becoming reliant on these devices to make behavioural decisions; suggesting that intuitive exercise was not practiced in these users. Given those with greater body appreciation tend to exercise intuitively [42], this may suggest a future intervention route to reduce the loss of body intuition. Furthermore, Ramsey [42] reported those engaging in the highest levels of exercise had low body appreciation, with a focus on weight and appearance, suggesting a fixation on health, and in this case WAT data, may be linked to low body image. While this needs further exploration, the current study did identify that some WAT users had an unhealthy relationship around exercise.

Given the reported trust participants placed in these devices and loss of bodily intuition, conflict was experienced when data mismatches arose. Frustration or upset was experienced as users were forced to acknowledge the trust placed in these devices, supporting user reactions within Dritsa and Houben [31].

Typically, a WAT’s judgement was adopted given their perceived expertise. For example, users reported feeling well rested until they viewed a low sleep score, which left them feeling tired. The self-fulfilling prophecy appears relevant here [43], suggesting users changed how they felt, and in turn how they behaved, depending on the data they received from their WAT. No research combining this theory with WAT use exists to our knowledge, highlighting an area for future research.

### 4.2 Study strengths, limitations, and implications

As with all research, the current study had its limitations. Despite containing incredibly rich data, the current study had a small participant size gained via convenience sample. Data saturation was reached during interviews meaning we would expect no themes to be introduced if further participants were recruited.

However, a different sampling method may have gained (less keen people coming forwards). The eligibility criteria of the current study was current, full-time WAT users. This sample allowed for in-depth insight to be gained around the novel topic of emotional conflict in WAT use, standing as the first study on this to our knowledge. Although, individuals who no longer use a WAT may provide a useful perspective in future research, particularly given that device cessation may have occurred due to emotional conflict arising.

Brands were varied in the current study, with four worn by participants (Garmin, Apple, Fitbit and Coros). This may reflect the popularity of wearables in the current UK climate, although, a wider range of WAT brands may have been beneficial for diverse experiences. Similarly, device heterogeneity occurred with all users wearing watch-based WATs. WATs in the form of rings or wrist straps without a display screen were not owned by these participants.

Given embodiment and dependence occurred from repeated on-screen notifications, user experiences around conflict may differ in these device types and should be considered in future literature. Finally, this cross-sectional study examined experience at one-time point, requiring users to reflect on complex experience in one sitting. Given emotions are ever-changing and conflict may arise and be resolved in very short durations, research examining conflict at repeated time points is recommended. For example, Ecological Momentary Assessment (EMA) has been used to gain health behaviour understanding and may allow for greater learnings on conflict experience and resolution over time [44,45].

The current study holds many valuable implications. A user profile emerged from data, with WAT users tending to be competitive, driven by numbers, and already focussed on health improvement. Given their perceived importance of a WAT in their health journey, guidance would be beneficial alongside the purchasing of a WAT around what constitutes as healthy use. For example, information around device accuracy and recommendations to consider provided data from a critical lens may benefit these users. WATs are beginning to be integrated into NHS healthcare settings as health promotion tools. The current study questions the introduction of these tools, with some WAT users facing conflict and negative emotion around device use. While physical effects appear to stand, the emotional toll identified in this study can question whether this is an ethical decision. WATs have been shown as effective tools for increasing activity levels and in turn weight loss [12], and therefore may be an excellent tool for some individuals, a patient’s personality and attitudes around health should be used to form judgement on whether WAT use will be emotionally beneficial. This recommendation is crucial given the long-term impact WATs may have on their users; many reported simply removing their device would not reverse negative emotional outcomes, with this knowledge gain and bodily awareness difficult to forget.

## 5. Conclusion

Emotional conflict was present in participants and presented within four themes; Who knows best?, Who’s in charge?, Who am I without it? and What’s happening to me?. While some WAT users navigated conflict rationally, others were strongly negatively impacted by this conflict. The current study highlighted potential dangers of WAT use, with the quantified self sometimes leading to a loss of bodily intuition in users. Participants reported perceived importance around their WAT and constant use, leading to WAT dependency.

Furthermore, WAT feedback seemed to promote physical activity, although this appeared to occur through feelings of pressure and have long-lasting impact on a user. The current study was the first to our knowledge on emotional conflict in WAT users, and holds crucial implications around device role out in healthcare services, recommendations around healthy use, and the long-term impact of using a WAT.

## Supporting information

Supplementary table of quotes

## Data Availability

Data will be made available via Figshare upon article acceptance. Alongside this, a full table of relevant quotes is provided as a supplementary document.

