## Supplementary table of quotes for "Tracking the tension: Examining emotional conflict experienced in wearable activity tracker users"

| Theme / sub-theme | Quotes |
| --- | --- |
| **Who knows best?**  This theme focusses on the perceived knowledge and expertise of a WAT, exploring how a WAT is used to define experience for many, what occurs when we disagree with the data received, and differences between how much trust we place in this information versus how we use it. | |
| The watch defines the experience | ‘When I didn’t wear a watch for a bit, remember when it was broken, well I still did it but I didn’t feel the same achievement on the amount of steps I’d done. I still walked the dog and literally did the same routes but not seeing a number meant it didn’t feel recorded. Recorded maybe isn’t the right word, but maybe acknowledged’ (JP).  ‘It somehow doesn’t feel the same even though I know it absolutely is. But I feel like I resent doing, say, going for a dog walk with it at home. Which sounds absolutely ridiculous to me’ (JP).  OPPOSITE: ‘I take it off when I go swimming each week even though some people buy these things to track swimming. In my mind it’s just, it’s not actually waterproof and I don’t know what I’d do with the fact I recorded a swim.’ (JP)  ‘If it dies and I can’t record my steps I find it dead annoying. It kind of feels like I’m not being rewarded for the work I’m doing’ (JP).  ‘I wear it to track all my workouts and you know in class you suggested would I, would I workout without a smartwatch on, and you saw me. I said absolutely not, there was no question about it… I wouldn’t workout without wearing one’ (MT).  ‘And how would I, in my mind, how would I know what I’ve done?’ (MT).  (With a watch) ‘I just know what I’ve done, like I know how long I was there, I know what my heart rate was, I know how many calories I’ve burned… I obviously would know like I went in at this time and came out at this time, but I wouldn’t know how hard I pushed and as much’ (MT).  ‘It gives me a score to beat instead of it being just on feeling or mood’ (MT).  ‘I wouldn’t know if I’d done 10,000 steps in a normal day without my watch, whereas if I haven’t, I’ll make sure I have with my watch, whereas I could have just been on like 8000 without a watch and not known’ (MT).  ‘I wouldn’t know if I’d met my targets without the data’ (MT).  ‘I remember thinking why would I take it off? I’m going to be dancing all night. I get to see what I’ve done!’ (MT).  ‘I just don’t know how I would know what I’d done in a day’ (MT).  ‘I just think that it would feel like it hadn’t counted’ (MT).  ‘I wouldn’t want to workout without a watch on because in my mind it doesn’t count. I’m still doing it but like, I don’t know, it’s not being recorded’ (MT).  ‘If it died during the day I’d be a bit miffed, a bit annoyed, or yeah like pretty angry if it was on a workout’ (AH).  ‘Oh I’m really bad for sleep scores, so my partner and I both have a Garmin and we’re terrors for judging how we sleep with that score. I laugh but it’s not good. So I’ll say morning and he’ll say morning, and then I’ll go how did you sleep and he’ll say yeah, 92, so good, and I’ll be like oh what I only got 81, like these random scores worked out from your watch’ (AH).  ‘I got -3 and I was fuming. I genuinely thought what’s the point? Should I fucking turn around? I was genuinely annoyed, not even because I felt strong, but more so because I’m on a run, isn’t that a good thing?’ (AH).  ‘I also think having a Fitbit started bringing back like disordered, not eating habits, but like mindset around exercise because the calorie count was just everywhere. So to compare with my Garmin, I’ve had a Garmin for about a year now and you actually have to seek out the calories you burn, it’s, it’s quite impressive, like you have to know where to look. They are absolutely not on your front screen … same for during a workout, during a workout you cannot see how many calories you’ve burned. You see your heart rate and, you can see the time you’ve been doing it, but on a Fitbit you could see the calories, it was actually like one of four statistics you saw when you were working out’ (AH).  ‘I really take it on board, I think I kind of listen to it more than myself which is pretty terrible’ (AH).  ‘If it’s like wah-wah you’re not doing good I think what’s the point in this run?’ (AH).  ‘I just fixated on it so the rest of it felt pointless, so I’ve sacked off workouts’ (AH).  ‘If I didn’t use it I would have no idea what kind of activity I was completing’ (HG).  ‘It frustrates me when I have to take if off and then I think well I can’t go anywhere now or do like any steps because they don’t count. It’s this whole thing over like, I can’t say I have’ (HG).  ‘Having the reminders on the watch and stuff when you already don’t feel well can be, like, demoralising. Thinking I’m not going to get my goal this week because I’ve unwell, and it’s saying you know, you’ve not done this and not done that. I think it can make me feel a bit like I’ve let myself down even though it’s like natural for people to need to take a break’ (HG).  ‘I think they need to have like an illness setting, you know? Something you can like put on it so that it knows that you’re not feeling as well’ (HG).  ‘You’re not going to go out and do the steps whether it tells you or not to at that point (depression), so I wish you had like a mood thing, you know, like, how are you feeling? And you could base the activity recommendation on what you’ve said your mood is for the day’ (HG).  ‘When you’re given so much data, then the idea of having that data taken away just makes you feel like how do I know what I’m doing? How do I know what I’m supposed to be doing?’ (HG).  ‘The whole thing is like a positive feedback loop, so it must have a positive impact on my head cos it’s, I use it as a means by which to validate what I’m doing’ (MH).  ‘Or I’m doing all that and my watch is telling me I’m literally not making any progress, so what’s the point?’ (MH).  ‘I probably couldn’t tell you above and beyond other than how ready to train I am what my HRV is gonna be, but stress is such a primal thing, ingrained in you as a human being. I don’t need a watch to tell me how stressed I am. It feels like it’s teaching me how to suck eggs, like I don’t need that’ (MH).  ‘I feel like it knows that better than me’ (MH).  ‘I was looking at the training readiness and training status, um to tell me like almost how I’m gonna do at the gym’ (ZA).  ‘I could just stop looking at that now, but my, my brain thinks like how would I know what I did?’ (ZA).  ‘The other day I had to charge it and we left the house and it was still on the charger and I said to my auntie, I was like I feel naked, I don’t have my watch on, and she was like how will you know if you’re alive?!’ (MN).  ‘I see people working out without tech and I’m like but what do you mean, like, like how do you know what you did?’ (MN).  ‘I could take it off but how would I know that I ran 5k today?’ (MN).  ‘I guess it comes back to that thing my auntie said, like, how will you know if you’re alive, how will you know your heart is beating? I’m like well obviously I know my heart is beating, like I’m alive, but it’s, it’s kind of comforting to just have it there’ (MN).  ‘I can look back and be like, oh, I actually have improved which is really helpful and also nice. Sometimes you feel like trash and you’re like oh actually I am better than I was two year ago’ (MN).  ‘It’s great I get this data but I don’t think I need that data much in reality, and actually having it, having it is probably, yeah it could be impacting my behaviour by having it, like I’m listening to it too much. I can just get out and do it and then I’ll judge how it was based on my experience instead of what a watch says’ (LP).  ‘I could go out on a big run in the hills and I’ve fixated on this number or distance, and without that number I’d feel good. I’ve had such a nice day. But then my watch gives me this judgment almost which, I know that’s then on me, but the watch gives me the information to me do strict’ (LP).  ‘That’s the issue. It feels, it feels important. It’s what I use to judge how I’ve done. Like it’s, I’m seeing it as this score that comes about everything else (LP).  ‘I’ll have to remind myself I’m doing it without the numbers still, I’m like doing it with or without data. The numbers record it but I’m still doing it. Cos at the moment I’m making a judgment only on that data … I’ve had to remind myself I do have a brain. I can make that judgement myself based on things like enjoyment and feeling strong and fast instead of, yeah, what my watch tells me I did’ (LP).  ‘It gives me a lot of statistics that I basically need in order to see if I’m improving or not’ (SA).  ‘The first thing me and my partner always do is “how did you sleep” and we both then look at our sleep on the watches and talk about it’ (SA).  ‘I do compare it to how it used to be and that’s how I see if I’m doing well or not’ (SA).  ‘I feel like if I’m not improving and I don’t have statistics and I’m… like what am I doing it for?’ (SA).  ‘It's the end of the week and you're like, oh, I've had a horrible week. And then you look back and you're like, actually, yeah, except looking all the times that I felt like happy or miserable, it actually kind of helps at the end of the week to think, oh, like, I haven't actually felt that awful all week. I've been OK. It's just right now I don’t feel good.’ (KL).  ‘I’ve never slept so good in my life, but it does come out of like, I know I'm only doing it because it's going to tell me I'm getting a good score in the morning kind of thing’ (KL).  ‘I'll purposely not wear my watch because I know I won't be getting home at least an hour. Do you know what I mean? So I just don't want it to know about that. I will purposely take it off just so my graph still looks nice’ (KL). |
| Navigating a data mismatch | ‘Say I wake up with 500 steps clocked. That can’t be true, but I just get excited and see it like a little bonus some mornings’ (JP).  ‘I’d question it, I’d like look into it and think about it… if I workout and it like seems a bit off I’d workout what happened’ (MT).  ‘If your average uni grade was like 70 and then you got 52 like you, you would question it. I think that’s pretty fair, pretty normal’ (MT).  ‘It felt like that workout felt harder but I think the watch is pretty good at picking up on things like illness and like my period. Like, I trust it probably felt harder for me than the watch is telling me and that means somethings going on with me. Like maybe I’m getting ill or slightly hungover still so it feels hard for me, but it’s less actual pay off’ (MT).  ‘There’s going to be a reason behind the numbers it’s pumping out and you get a watch to learn more, so I’m not going to argue with it. Like I’ll try work things out, but I’m not going to argue with it and tell it it’s wrong when it’s a tool, like, it’s tracking me. So I’ll just think about why it might feel harder for me’ (MT).  ‘I started getting really frustrated at it because there was such a lag in data when working out. I have a spin bike at home and I was using it and I was just looking at my heart rate and like I would be going at my absolute max and it would just keep glitching, saying my heart rate was 90, like 90 beats per minute, and then I would stop to try reset it and it would say like 170 bpm, which I know doesn’t effect the workout over the effort you’re putting in but oh my god it ruined it for me, I would get so frustrated and I actually stopped half way through a couple of times’ (AH).  ‘It’s really, almost been the rational information I’ve needed when my brain has been devilish and on fire and really quite harsh, it’s given me that objective information’ (AH).  ‘It was just completely wrong and actually that sent me into such a spiral. I questioned like what if I’m running at insert a really slow number, what if I think I’m alright at this and it turns out all that data was wrong? I’m actually not fast at all? So that was a real spiral. I waited maybe 3 months, I didn’t go running in that three months apart from on a treadmill so I could see the actual data. How mad is that? I just didn’t want to, it just really weirdly knocked my confidence’ (AH).  ‘By day five it told me I was overreaching so it genuinely informed me to have a quiet weekend. Even though I saw it and I was annoyed, and I was a bit like nah, I’m good, I trust it kind of knows what it’s talking about so I didn’t do anything on Sunday’ (AH).  ‘Even though I could have had the most well rested sleep ever, I could feel incredible, it actually happened the other day. I had such a deep sleep and it gave me 70, and I was like almost disappointed. But I also never know if then I see that score and it unconsciously makes me feel more or less tired’ (AH).  ‘I was so frustrated with my Fitbit because it just wasn’t accurate, and I know you shouldn’t put all your faith in these things, but I think I do a little bit’ (AH).  ‘With my Fitbit I got furious from the lack of agreement there’ (AH).  ‘I suppose there’s that frustration as well that I mentioned, like no I feel fitter than that or yeah just like I don’t actually think this is correct, but then at the same time I completely trust the watch, so you know, I’m more likely to disagree if it’s bad news. If you don’t feel very fit and it tells you you’re amazing, you don’t really question that as much do you, because, oh, what a lovely compliment! So I never know if it’s a little bit of defiance’ (AH).  ‘I might feel well rested and then I’ll see I got a 62 out of 100 and I might then feel tired’ (AH).  ‘I swapped from a Fitbit because the data was, I realised it was so inaccurate but I was taking it as the truth, I really got quite, look I’m going to use the word distressed. I was not in a good way after that realisation’ (AH).  ‘If I disagree with it, I kind of take the watches views over my own. When I say it out loud it doesn’t sound good’ (AH).  ‘I really take it on board, I think I kind of listen to it more than myself which is pretty terrible’ (AH).  ‘I’m just about to contradict myself because I was going to say I feel like you just remind yourself it’s a watch every now and then, but also when I found out it was giving me wrong running paces I didn’t have a balanced mind, I cried with anger’ (AH).  ‘I really don’t like, it’s when you do like a workout or like a walk or something, it gives you an effort level, but it’s not tailored, you don’t have any say… it’ll just naturally say like easy effort and it’s kind of offensive because you don’t know who I am and you don’t know… I hate that’ (HG).  ‘For example, I take my dog out for a walk so I could be walking quite quickly but because he wants to stop and sniff it just takes the pace down, and it’s saying your heart rate is too high for your speed now’ (HG).  ‘It would be better to like, you know, ask people more personalised questions about how people find exercise and like what their targets are before you just slap an effort score on because oh my god that was high effort for me and it’s pretty rude’ (HG).  ‘I know my body better than that’ (HG).  ‘It’s taken me a while to like, you know, take it with a pinch of salt because when you start trying to exercise more and stuff, you rely on it because you don’t know any better’ (HG).  ‘It just doesn’t sync to like life events very well, does it? It’s just expecting every day is going to be the same, like you can get up and go out every day and you can’t’ (HG).  ‘I sort of try and take what it tells me with a pinch of salt if I’m not feeling that. I use it more to validate how I feel rather than have it tell me how I should feel… I will go to it for validation rather than it be the source of truth. It’s almost like a second opinion for me’ (MH).  ‘I feel like say four years ago when it was like new to me I’d rip my hair out, there was an element of frustration for sure. I’ve got all these resources telling me not to push too hard but I feel good. Or I’m doing all that and my watch is telling me I’m literally not making any progress, so what’s the point?’ (MH).  ‘[Exercise is] very honest and you can’t lie about how it went, that’s just how you perceived it, so I feel like the feedback from the watch is just misaligned to your feelings but [the watch] is so much more concrete’ (MH).  ‘I would lean towards technology in that like I, I feel like as a person you’re only capable of considering so many variables at one time, whereas these, the whole point of technology is that it looks at things much more holistically. It’s got the full picture and it’s privy to, considering variables that you’re not necessarily even aware of’ (MH).  ‘I think I would always lean to trusting the watch in a situation like that. My behaviour might not respond because, say, I feel a bit scatty and want to go on a run, but I’d trust my watch still’ (MH).  ‘I get a bit confused if like I’m pushing really hard and it shows my heart rate is really low, yeah, I think I’ve done a good workout and it’s just not showing that. Then I’m like, I try to unpick why i’ve got the data back but I don’t agree with it. To, to be honest, I just tell myself that I’m sulking cause I don’t like the feedback. That’s probably right so that’s what I go with, because you wouldn’t question the data if it was a bit nicer than you expected would you?’ (ZA).  ‘It could say my heart rate has hardly moved or I got a slow time which I just find pretty [pause] I don’t know, I see it and I think ‘huh’ it didn’t feel like that. I think I just kind of shrug it off, and I, see if I’m lifting weights it’s different because I know the reps i’ve done, but if I’m running cos that’s like, I can’t measure a 5k and I’m not counting, so if I’m running I’ll think okay maybe I wasn’t as fit as I felt that day or maybe I didn’t push as hard as I thought, or I did but things like the fact I’m tired or or it’s hot, erm yeah, that’s when I go with the watch’ (ZA).  ‘It’s more accurate than me, like I can’t to 28 minutes of something, so I think you’ve got to [go with the WAT] there’ (ZA).  ‘Sometimes it says I’m strained when I feel strong or the other way round. That one I find annoying, really annoying if I feel strong. I think annoyed is fair, just frustrated about it because I feel good and it’s telling me I’m not’ (ZA).  ‘I’m like really interested in my heart rate, like how it perceives my recovery and then if that matches up with what I’m feeling because I will always go with how my body’s feeling rather then the watch, which is something that I’ve had to develop over time and not have the tech rule my life… maybe that’s an age thing’ (MN).  ‘I’m more guided by what my body is doing rather than what the watch is telling me at this point in my life. It would have been, a past time where I was like, if it said like one of the predicted activities was [outside of what I could do] I probably still would have gone and done it’ (MN).  ‘Feeling really good in your body and then coming back and your watch is like oh you ran really slowly… now I don’t know. Like it kind of makes me second guess. Not like completely rewrite how I feel, but absolutely question it’ (MN).  ‘Sometimes I can be like oh whatever, and then other times depending on what my mood is, it probably affects me more negatively’ (MN).  ‘I do get frustrated at it and I think that sometimes affects the rest of the day’ (MN).  ‘I don’t know how to explain that so I kind of just ignored it, and I was like well I know I ran this time, the watch clearly doesn’t, but that does annoy me… that just confused me too. Honestly, I think confusion was the main one because I do trust it’ (MN).  ‘Sometimes I agree with it but there’s definitely other times when I have been surprised to see its feedback, but I’ve still listened to it’ (LP).  ‘If I see I’m not ready to do something, even if I feel okay, it just seems I’m a bit strained, I get quite stressed’ (LP).  ‘I could feel really well rested but I see a sleep score and it’s just like I have a full debate, I just think oh well I must have actually slept barely’ (LP).  ‘I use that stress app that tells you, oh, you’ve got a raised heart rate, stress levels. Don't know what I'm stressing about, but we need to like take a minute and but then at the same time it is, it could be negative because it is telling me that I'm stressed when I'm actually feel quite all right. That's kind of the problem is you you'll be getting on just like doing regular stuff and then it'll tell you you're stressing. You're like, right. Well, now I'm even more stressed because I don't know what to do about it now.’ (KL).  ‘It's the end of the week and you're like, oh, I've had a horrible week. And then you look back and you're like, actually, yeah, except looking all the times that I felt like happy or miserable, it actually kind of helps at the end of the week to think, oh, like, I haven't actually felt that awful all week. I've been OK. It's just right now I don’t feel good.’ (KL). |
| **Who’s in-charge?**  This theme focusses on the determiner of behaviour, examining the impact a WAT has on user behaviour, but also questioning whether this impact routes back to the user themselves. | |
| The user behind the device | ‘I suppose getting older and sometimes being ill makes me feel pretty uncomfortable already so I could be blaming it for my own feelings’ (JP).  ‘I got one when I was already so deep into exercise’ (MT).  ‘It gives me a score to beat instead of just on feeling or mood’ (MT).  ‘I’ll be like, you reached like 300 calories at this point last time and you’re on 250 so you can push more’ (MT).  ‘I’m just pretty competitive with myself and it gives me a way to have that’ (MT).  ‘I don’t know if that’s the watch or me with like exercise as a whole’ (MT).  ‘Do the numbers make me stressed though, because if I did, if I worked out without the watch on I’d be even more stressed’ (MT).  ‘I know it was actually my fault as a user but it got to like this weird count up of how many calories you burned, whereas with a Garmin it’s like look I’m trusting you, do your workout, and then you can find out after if you really want to know, but again, it’s not shoved in your face’ (AH).  ‘The bit I actually cared about and went in to check first was calories for sure, and that’s just because of the mindset I was in’ (AH).  ‘I just really like to see the numbers tally up’ (AH).  ‘I can’t not see the notification of the score when it pings up but I have quite a distracted brain so that is probably on me’ (AH).  ‘It’s hard though cos I think that pressure is like internal, but it’s pressure from the knowing of the watch. I think what I’m trying to say is that someone else could get that data and do nothing with it, and it’s me being quite anal and single focussed that means the data has that effect on me’ (MH).  ‘It’s odd to say (dependence), but I also feel like I made the purchase for that reason’ (MH).  ‘It does impact my behaviour a lot. That’s the point of a watch for me though’ (ZA).  ‘I think it makes me more competitive with those numbers’ (ZA).  ‘I really like seeing my scores go up’ (ZA).  ‘I was chatting to a friend and we said it was a bit like, do you remember doomscrolling in COVID? It was a bit like doomscrolling and fearmongering. It can feel a bit like that, like I’m, i’m almost actively looking at something to stress me out’ (ZA).  ‘I wanna say motivation because it sounds better doesn’t it? I wanna say motivation but I think it is pressure a bit. I think it’s hard to know if I’ve put that pressure on myself or the watch, maybe I’ve put pressure on myself to stick to these things…I wanna say I’m putting pressure on myself but then hm, let me think, my watch provides the data that I can then use to pressure myself’ (ZA).  ‘It makes me a little bit stressed but I think that’s just cause I’m really locked in at the moment’ (ZA).  ‘I think I just know a bit too much from it, but also that’s what I like and why I got it’ (ZA).  ‘I have come from a background of exercise addiction so I’m very careful on trying to not slip back into that so that’s one way I kind of take the pressure off myself (removing calories)… but then like, is that the watch doing that or is that just my personality? Like can be blame the watch?’ (MN).  ‘I use V02 max with a grain of salt as well, but I get really excited when that goes up’ (MN).  ‘I also love the time predictors for a race because I endurance run so I, the predictions, I like to beat them’ (MN).  I don’t know if there are any positive emotions except when you PR and it’s like woo well done! But maybe the chasing of the feeling is enough’ (MN).  ‘I set myself lots of goals anyway. Love a goal. I’ve always been white goal driven and the watch means my goals are based on numbers’ (LP).  ‘I know that’s then on me, but the watch gives me the numbers to be so strict’ (LP).  ‘Instead of you keeping track of what you’re doing there’s a device that does it for you it’s quite nice [because] I used to have like a wellness journal of what I was doing’ (SA). |
| Guided or governed behaviour | ‘My children use them as second phones and to me it looks like the watch makes decisions for them’ (JP).  ‘I can be listening to the watch more than myself’ (JP).  ‘I wouldn’t change my plans in a large way, but if I’ve not done much I could, say, park far away from the shops and go that longer route’ (JP).  ‘I get a notification every so often telling me to move and sometimes I do move from that. It’s never far but if I want a break from whatever I’m doing or if I agree that I’ve been stuck in the same place I’ll go for a little wander, maybe get water or something’ (JP).  ‘Oh god, [the data] is so important. I use it. I just, I just use it all the time. I use it for my eating. I use it to decide if I should walk to the shops. I, I use it to plan, like, I think because it’s, I have my daily targets to meet it’s well important’ (MT).  ‘I use it to plan should I go for a walk or should I sit on the sofa, or yeah, like what meal to eat that night. I look at the calories I’ve got left and yeah, I’d have it planned like that, so yeah, the data is the reason I got the watch’ (MT).  ‘I look at my move goal like weekly, so it gives you like a Sunday summary and I look at that to see when I need to put more effort into moving a bit more. So this is on, on Wednesday and Thursday I didn’t get my move goal so I know what maybe like this week, Wednesday and Thursday, I’ll use my walking pad when I’m working’ (HG).  ‘I try pick out the reasons why I want to do something. Is it for me or is it for the watch? Sometimes I’m not sure’ (AH).  ‘I look at it and rely on it, and use some of the data to dictate what I do’ (MH).  ‘I did that yesterday because I noticed it was low (step count) given I was working from home all day so I walked to the shop instead of driving’ (MH).  ‘I definitely feel like I weigh it up more from the performance and calories side of things. [Drinking alcohol] is not good and I definitely learned just how bad it was from my watch’ (ZA).  ‘I mainly use it for essentially knowing what I’m doing each day’ (ZA).  ‘It can predict the calories you’re going to burn by the end of the day and then I use that to see what I can have. I’m currently on a cut so I will use that to inform if I’m still hungry what I can have’ (ZA).  ‘I do look at the others a lot and they will kind of, kind of inform my decisions a bit… I don’t like saying I look at my watch to know what to eat, but it’s also calories in calories out. It is really useful for that, but that’s, that’s the type of, the type of thing I say and think that doesn’t sound good’ (ZA).  ‘It gives a bit of tough love in the fact that if I want to eat more I need to move more’ (ZA).  ‘It’s been chucking it down and I’ve not wanted to go for a walk in it, but I have to get steps in. Which to me, that’s discipline, but my girlfriend is like well don’t do it then, and I think it comes from the pressure of wearing a watch. Or like, if I’ve not moved, and I’ve not been active, so I’m like, like doing steps around my kitchen. Not madly pacing or anything, but I’m trying to talk about more, to get steps up. But my steps come from my watch so that’s probably pressure, pressure yeah, but that makes me feel a bit weird’ (ZA)’  ‘I do think, you know when I mentioned toxic, I do think my girlfriend is right a bit of the time, like we had a massive lie in the other say. She’d been out and I hadn’t, and she was like what do you keep checking on your watch? And I was checking the time because it was like 2:00pm and we’d not done anything and my step count was basically zero, so I was thinking yeah I probably need to move soon’ (ZA).  ‘Well it’s what I use it for, it’s why I got it so it’s really important…I use the data, well, I use it to like decide what I can do’ (ZA).  ‘I’m just a bit aware that because I’m in a cut right now it doesn’t sound healthy at all, but yeah, I use the watch to decide that and to let me know what I can have’ (ZA).  ‘It might sometimes make me exercise or do stuff when I don’t actually feel like it or when my body doesn’t really want to, but I’m like, oh, I’ve got to do it’ (MN).  ‘I’ve debated stopping wearing one because I find it’s just like I’m listening to it a bit too much’ (LP).  ‘If I hadn’t slept well, would wake up and use it to justify not going on a run, whereas if I didn’t know that I would have thought I didn’t sleep the best but didn’t sleep the worst. I would have still gone on my run and chances are I would have enjoyed it, and so the reason I’ve been debating stopping wearing one is because I feel like I have been listening to it a bit too much’ (LP).  ‘I’ve almost lost like my own autonomy with it’ (LP).  ‘It will tell me like how ready I am and how fit I am to work out and I do find myself listening to that a lot’ (LP).  ‘That’s where I’m listening to it a bit too much. I think the thing I’m struggling with is that you get a smartwatch to change your behaviour. Generally I got a smartwatch because I thought it would help me be fitter, and so it is changing me in that sense, but now, I don’t know if this comes with getting older and unpicking it, but now I’m questioning like if a watch should be changing my behaviour. It definitely changes my behaviour, I just don’t know how I feel about it’ (LP).  ‘It says I’m maintaining training status so I guess I need to do another cycle ride today’ (SA).  ‘Tonight you will have to go to bed at 8pm so I also, it does govern a little bit of my sleeping schedule as well’ (SA).  ‘With your watch, you would sort of know that as you will plan your exercise based on that, you'll be like, okay, well, my, like, for example, I know for a fact that I have not slept well six hours and I know at a certain point today I'm going to have a crash. So then I'm scheduling my cycle race like earlier than rather than later’ (SA).  ‘I do think it's helped my sleep 'cause it almost gives you like the push you need. I’ll be like, right, If I go to sleep now my Apple Watch will tell me I'm doing good in the morning.’ (KL).  ‘It's like a little tyrant on my wrist. And I'm forgetting that I've got free will and can take it off’ (KL).  ‘I'm getting controlled by a watch is basically what I'm telling you’ (KL).  ‘I'll purposely not wear my watch because I know I won't be getting home at least an hour. Do you know what I mean? So I just don't want it to know about that. I will purposely take it off just so my graph still looks nice’ (KL). |
| **Who am I without it?**  This theme focusses on the dependence on devices some users face, exploring the concept of device embodiment amongst participants, a fear of the potential removal of this device, and the lack of clarity in what impact this removal would have on behaviour. | |
| Device embodiment | ‘I put it on and was instantly like oh my god this has changed everything’ (MT).  ‘I’ll just use it as a second phone. So like I think that’s why I love it, because it’s apple everything’s just like a second phone on it. So I get my messages on there, I get my notifications on it, it’s it’s just an extra screen so yeah, kind of everything’ (MT).  ‘I do debate if I’m dependent on it, which I think most people do with technology’ (AH).  ‘I try pick out the reasons why I want to do something. Is it for me or is it for the watch? Sometimes I’m not sure’ (AH).  ‘I’ve never stopped using a smartwatch since I got it, like it has genuinely stayed on my wrist the whole time. The extent for debating [if to not wear one] is going ooh that’d be weird’ (AH).  ‘I use a dexcom blood sugar monitor and its integrated into my watch so I have a graph on the watch that shows me my blood sugar and I’ve got an alarm. It will come up on my watch, so I don’t have to get my phone out’ (HG).  ‘When it’s on my wrist and it’s so close I feel like I can’t separate it from myself’ (HG).  ‘It’s weird… it’s really difficult to separate between what is me and what is the watch’ (MH).  ‘I’m just pretty into it, I’m on it a lot and wear it all the time’ (ZA).  ‘It’s hard to know if I’ve put that pressure on myself or the watch’ (ZA).  ‘I wouldn’t stop using one so I can’t imagine it’ (ZA).  ‘The other day I had to charge it and we left the house and it was still on the charger and I said to my auntie, I was like I feel naked, I don’t have my watch on, and she was like how will you know if you’re alive?!’ (MN).  ‘It just goes everywhere with me’ (MN).  ‘I’m not sure where it ends’ (LP).  ‘I’m thinking about seeing how I feel when I take it off, I think it will make me feel a bit strange for a while, I will forget I’m not wearing it and go to check it because it’s just a part of me’ (LP).  ‘It sounds stupid but the first thing that popped into my head was that it might be boring, it might be a bit lonely, because I really like waking up and seeing what I’ve done. It’s like a paper in bed to me and it’s every day and always there at the moment, so yeah’ (LP).  ‘I think if you’ve worn it for so many years as well, it’s normal, it’s a big routine you’ve formed. It’s the normal now so it’ll take a lot of reversing for sure’ (LP).  ‘I think I'll be actually lost without it’ (SA).  ‘I feel a lot of it is habitual. So like, I check it however many times a day. A lot. Because I’m so used to having it. It can be dead and I’ll still go to check it’ (CF). ‘I've got diabetes, so it'll tell me like when I need to when I need to change my Dexcom and everything, so I use that setting quite a lot because I'm very good at forgetting and I have to not forget. I need it!’ (KL).  ‘It's like a little tyrant on my wrist. And I'm forgetting that I've got free will and can take it off’ (KL). |
| Fear over device removal | ‘I wear it to track all my workouts and you know in class you suggested would I, would I workout without a smartwatch on, and you saw me. I said absolutely not, there was no question about it… I wouldn’t workout without wearing one’ (MT). ‘You said would I workout without once a week without a smart watch and I just wouldn’t… or if it broke I would still do it but I’d hate it. Like even now it just, you can tell I’m just so stressed. It feels, it stresses me out even imagining it’ (MT).  ‘If I worked out without the watch on I’d be even more stressed’ (MT).  ‘It’s not normal. I shouldn’t have that reaction to, to working out without it, but I, I’m just not doing that. Like it makes me feel sick, it’s not normal, is it?’ (MT).  ‘I do debate if I’m dependent on it, which I think most people do with technology cause I’d probably panic if I didn’t have it’ (AH).  ‘If I was told to take it off for a month I would definitely panic, erm, which I don’t think is good. I’m still doing the same thing even if it’s on or off my wrist, but it reassures me, I really like it’ (AH).  ‘I almost hypothesise what would happen and then I feel panicked’ (AH).  ‘The specific panic I feel comes from if I took one off would I gain loads of weight? (AH).  ‘One part of me goes oh that would be so interesting to do, I should try that, and the other part of me in the exact same moment goes shut up, absolutely not, I can’t think of anything worse. Yeah I don’t know what my reaction would be and it’d be pretty interesting, but I’d definitely be panicked’ (AH).  ‘It feel like its kind of obsessive at this point, like not just me, but other people, because like even like you say, if you didn’t wear it, the activity makes me uncomfortable. That question makes me uncomfortable’ (HG).  ‘When you’re given so much data, then the idea of having that data taken away just makes you feel like how do I know what I’m doing? How do I know what I’m supposed to be doing?’ (HG).  ‘I didn’t realise how obsessed I was with it until you said like about taking it off. I was like I don’t want to do that!’ (HG).  ‘I had to wait like a month or so to replace it, so I was completely free, which at the beginning made me feel really anxious’ (MN).  ‘I could take it off but how would I know that I ran 5k today?’ (MN).  ‘I guess it comes back to that thing my auntie said, like, how will you know if you’re alive, how will you know your heart is beating? I’m like well obviously I know my heart is beating, like I’m alive, but it’s, it’s kind of comforting to just have it there’ (MN).  ‘I think I’ve become quite anxious is I’m not, like if I woke up tomorrow and it was broken, first of all I’d be made, but but yeah I think I would be anxious if I didn’t have something there to track, which saying out loud sounds really silly, but yeah, I think the anxiety of not having it, but also acknowledging the idea that it does make me anxious sometimes. It literally makes me anxious but the idea of not wearing it also freaks me out’ (MN).  ‘If I didn’t wear it, then it wouldn’t be the end of the world literally, but I would hate it…I don’t wanna go without it’ (MN).  ‘I would cry because I wouldn’t have a reference point if I’m not doing well or not’ (SA). |
| **What’s happening to me?**  Finally, this theme focusses on the outcomes of wearing a WAT, focussing on the mechanisms of this behaviour promotion, the belief that perceived success comes alongside a compromise in emotions, and the impact having access to this information had on participants. | |
| Mechanisms of behaviour | ‘I think it is motivation because I like seeing that number go up. That feels good and like an achievement. I’ll even say to my partner wow look at my steps and that implies it’s an achievement, and I’m driven to do a lot of steps because it’s good for you. Motivation is the front emotion for me, but I do have to recognise that I like getting steps in because I know they’re good for me, so I feel like there must be a bit of pressure to do that’ (JP).  ‘I’m a bit out of sight out of mind as a person, and, and I’m nearly 60. I know I’m not in my prime so the data probably wouldn’t make me feel good. Not that I’m fully avoiding it, but I know I’m a little ropey now!’ (JP).  ‘That depends on how I’m doing, you know, if it’s an active day or not. If it is, I feel great and it’s an extra pat on the back, but if I’m having to have a slow say it feels like a reminder that I’m not doing great. Yeah, it’s a double edged sword that can be uplifting or shaming depending on what you’ve managed’ (JP).  ‘That’ll make me feel a little down, if I’ve not done a lot that week’ (JP).  ‘I want to say motivation because like the numbers really motivate me. I love seeing the numbers go up and I know high numbers are good for me, but because I said that like not wearing my smartwatch would stress me out so much, I don’t really know how to unpack it, I think it’s the numbers. I think it’s motivation but, well, I can see it’s unhealthy’ (MT).  ‘I get anxious thinking about, so if I’m looking at my watch and I’m looking at the time, I’m thinking you’ve not worked out yet and you’ve only done 4000 steps’ (MT).  ‘The numbers motivate me to do it, but it’s out of stress’ (MT).  “If I’m not doing it I feel pressure, so yeah, I don’t, it’s kind of both’ (MT).  ‘It’s really hard to unpick if it’s cause I want to do it or if it’s cause I feel like I need to/ I feel like I need to do it even though I know like if I don’t go to the gym for a day the world won’t end. But I do get stressed at the idea of not’ (MT).  ‘The numbers being low stress me out’ (MT).  ‘It motivates me but I know that it’s like stress as well, like if I’m doing bad, say if I’m poorly, I know I wouldn’t be reminded I was doing bad without a smartwatch, whereas I’m wearing one and I feel bad about it telling me I’m not doing anything. Yeah, it can stress me out’ (MT).  ‘I can see my heart rate and calories during working out and so I think from that, that makes me competitive’ (MT).  ‘It makes me feel really good when I see the numbers when I’ve met my goals, but if I’ve not met my goals yet, it makes me feel really stressed because it’s just this, like, flashing deadline’ (MT).  ‘Having the data on the watch makes me feel good. It like says look and that you’ve done!’ (MT).  ‘I got -3 and I was fuming. I genuinely though what’s the point? Should I fucking turn around? I was genuinely annoyed, not even because I felt strong, but more so because I’m on a run, isn’t that a good thing? I felt like I was being punished for going on a run’ (AH).  ‘I, generally, deep down do not believe I’m good at sport. I don’t believe I can be good at sports and that’s from some horrible PE lessons, and now I’m trying to unlearn that, and so for example, I go on a run and I double take at the speed I’m doing. I almost don’t believe it and I know for a fact without that watch I wouldn’t feel as strong and healthy, then empowered to do more’ (AH).  ‘It’s… can I say both? I think sometimes it motivates me, it probably motivates me if I’m already in the flow, so yeah, let’s keep this up, or yeah you’ve done 8,000 steps you can get to 10,000 by this time, nice. I feel like it can work the other way though and especially from in the bad way, maybe it depends on my mindset and where my head is cause if I see I’ve done something like 200 steps, or I’ve not worked out in a week and race predictors is increasing and my like training status is dropping, that’s when it could be… not pressure, but guilt. I can feel a bit guilty, a little bit down in the dumps’ (AH).  ‘I literally think it’s both. It could almost be like it pressures me to get out but once I’ve got out, it means I’m more empowered. It like on one side it says get up lazy, but then the data on the other side says look at what you did’ (AH).  ‘It had caused me to feel a little bit hmm I wanna say anxious but it’s not anxious, maybe like a little bit on edge? I think that just comes from being aware of your movement when you start to care about something, like if there’s a random Sunday and you’ve only done 2000 steps and it’s nearly evening you’re aware of that. I don’t want to start like marching around… you just have that awareness and sometimes that’s really empowering but other times like it’s not a switch off’ (AH).  ‘I try pick out the reasons why I want to do something. Is it for me or is it for the watch? Sometimes I’m not sure’ (AH).  ‘It does make me like happy, I do like to use it, but sometimes I do find it stressful. Like if I’m a bit tired and it’s going off all the time, it can be, yeah’ (HG).  ‘It depends like if I’m feeling motivated already, it like helps that because I can see my progress being reflected back to me, but if I’m unwell… having the reminders on the watch and stuff when you already don’t feel well can be, like, demoralising. Thinking I’m not going to get my goal this week because I’ve unwell, and it’s saying you know, you’ve not done this and not done that. I think it can make me feel a bit like I’ve let myself down even though it’s like natural for people to need to take a break’ (HG).  ‘It was irritating and it’s like, yeah, disappointing… I would just take it off and just ignore it because it was starting to bother me too much’ (HG).  ‘When I’m feeling good and like motivated, the reminders and stuff do help, and like tracking steps, it’s more motivation for me to get there because it says you are, you’ve met like three quarters of the progress, you’ve only got this bit left and that is really motivating’ (HG).  ‘It’s like totally up or totally down depending on your mood, like, if I’m doing well it can be really motivating, but if you’re not, well, it can just be the total opposite and very what’s the point?’ (HG).  ‘I really don’t like, it’s when you do like a workout or like a walk or something, it gives you an effort level, but it’s not tailored, you don’t have any say… it’ll just naturally say like easy effort and it’s kind of offensive because you don’t know who I am and you don’t know… I hate that’ (HG).  ‘I do just love data, so I do think that being able to get into the granularity of a session after a session is like a feedback loop. So it’s, it hasn’t motivated me to do the thing, but I’m a little rat who gets more cocaine at the end of a workout because I’ve got this like, it’s like I do the exercise then I get this endorphins kick after exercise, but then also three hours later when I’m sat in the bath I can look at the numbers and get an extra pat on the back while I fine tune going forward’ (MH).  ‘I feel like getting into stripping back the layers of what that is, I can say it’s because I like looking at the numbers, but at the end of the day it is just a different form of gratification’ (MH).  ‘The whole thing is like a positive feedback loop, so it must have a positive impact on my head cos it’s, I use it as a means by which to validate what I’m doing’ (MH).  ‘I can see stuff went as well as I thought it did and that feeds back into me wanting to continue doing it’ (MH).  ‘Coming back to that ignorance is bliss, I definitely think wearing a smartwatch has had an impact on my relationship with alcohol, in that, I feel like I’m now so aware of the impact it has on my sleep and overall training readiness and I can’t but the cat back in the bag now with that. So I feel it’s probably created quite a bit of pressure to not drink there’ (MH).  ‘I have found sometimes where I’ve not drank and then I’ve had to say to myself oh come on, you’re 29 and you actually want to drink but you’re feeling told off by a watch’ (MH).  ‘It’s not guilt it’s just like awareness and you can’t un-know something, so more that pressure from the information’ (MH).  ‘It can feel a bit like that, like I’m, I’m almost actively looking at something to stress me out’ (ZA).  ‘I wanna say motivation because it sounds better doesn’t it? I wanna say motivation but I think it is pressure a bit. I think it’s hard to know if I’ve put that pressure on myself or the watch, maybe I’ve put pressure on myself to stick to these things…I wanna say I’m putting pressure on myself but then, hm, let me think, my watch provides the data that I can then use to pressure myself’ (ZA).  ‘I think over time from seeing the data and how it changes when I’m more active, it’s motivating me over time, but it’s probably come from the fact I feel pressured to stick to it. Like now I know, now I know how much movement actually impacts the calories burned and what my body needs it could be pressure’ (ZA).  ‘It’s been chucking it down and I’ve not wanted to go for a walk in it, but I have to get steps in. Which to me, that’s discipline, but my girlfriend is like well don’t do it then, and I think it comes from the pressure of wearing a watch. Or like, if I’ve not moved, and I’ve not been active, so I’m like, like doing steps around my kitchen. Not madly pacing or anything, but I’m trying to talk about more, to get steps up. But my steps come from my watch so that’s probably pressure, pressure yeah, but that makes me feel a bit weird’ (ZA)’  ‘I do think, you know when I mentioned toxic, I do think my girlfriend is right a bit of the time, like we had a massive lie in the other say. She’d been out and I hadn’t, and she was like what do you keep checking on your watch? And I was checking the time because it was like 2:00pm and we’d not done anything and my step count was basically zero, so I was thinking yeah I probably need to move soon. So that’s, that’s when she was like that’s so toxic. I think it makes me quite anxious having the data, but it’s the anxiety that brings, is what makes me do it, so it’s kind of like a mean teacher that I don’t like, but then I’ve opened my results and think you know what, they knew what they were doing’ (ZA).  ‘I’m just pretty into it, I’m on it a lot and wear it all the time. I really enjoy looking at the data but I can sometimes find myself panicked on numbers like heart rate and steps when nothing has actually changed in the last year, I just have the information now’ (ZA).  ‘It makes me feel that sense of achievement like I do when I’ve had a really good day. By good I probably mean really active…but if I’ve had a bad day then I’m anxious, so it basically depends on what the feedback is. It gives me that feedback whether I want it or not and if the feedback is good or bad depends on if I like the watch’ (ZA).  ‘My step goals really low on purpose so I don’t stress about it’ (MN).  ‘There are some days when I’m like oh, none of this matters, and then the next day I’m like oh why was my heart rate so high?’ (MN).  ‘I focus on it more when I’m actively training for something’ (MN).  ‘I also find the Garmin langue really kind of negative in terms of like at the end of your run or whatever it’s like this was productive or unproductive whereas apple will say oh well done, you went for a run!’ (MN).  ‘I do get frustrated at it and I think that sometimes affects the rest of the day, mood wise, I think yeah, definitely. It stays on my mind, it takes away from the fact I’ve done something really good’ (MN).  ‘If it tells me I’m really ready to go, I’m motivated, right? I’m like yes, nice, I’m ready. But if it tells me I’m really tired and I shouldn’t go out, I think because I’ve been injured in the past… if I see I’m not ready to do something, even if I feel okay, if it just seems like I’m a bit strained I get quite stressed’ (LP).  ‘If it tells me I’ve done well, I feel good, stating the obvious right?’ (LP).  ‘I think because the data dates me really competitive in quite an anxious way [removing the WAT] will make me more relaxed’ (LP).  ‘I panic when sometimes I look at my training status and it says something like maintaining, because then I think oh so, like that’s not good because it usually says optimal’ (SA).  ‘I do feel guilty in the sense, if I don’t do 8,000 steps I do feel guilty, but not because… I feel guilty that I haven’t invested in my health on that day’ (SA).  ‘I used to get quite excited about the VO2 when that used to change because sometimes it says like you’re above your age, and it gives you like a, you’re like a 32 year old so I’m like yes! It does motivate me’ (SA).  ‘I noticed a difference and yeah, I find it quite motivating. It's a big motivator for me’ (SA).  ‘I know that they found it quite disheartening to know like they were doing like, let's say, 2000, 3000 steps a day, and they couldn't do 8000. And the fit people was letting them know that they were sort of losing, you know, like the fitness. So I can understand that if you have a chronic condition, it might not be the best, but if you are healthy it can do so much good’ (SA).  ‘It does help me because you get such a boost because you feel like you’re doing well, then you invest more time into it’ (SA).  ‘I tend to [take it off when I shower] just because I’m worried my average heart rate on the day will go up’ (SA).  ‘The goals you set with like the activity make me more motivated when I want to reach the goals I’ve set’ (CF).  ‘I don’t think there’s any pressure, like if I don’t close my rings one do I’ll do it another day. It doesn’t really bother me that much but I feel I mainly use it for notifications’ (CF).  ‘If I’ve closed [the rings] I’m happy, but if not that’s fine’ (CF).  ‘I do think it's helped my sleep 'cause it almost gives you like the push you need. I’ll be like, right, If I go to sleep now my Apple Watch will tell me I'm doing good in the morning.’ (KL). ‘I'd say it's probably mixed. It depends when it's telling me to. Last night, perfect example, I had literally just got into bed and it was like there's still more time. Close your loop and I was, oh, I cannot tell you how quickly I swiped that notification away. So times like that, absolutely not like. But if I've kind of been sat for a while, like if I've been sat doing my uni work, but like, not doing something active for a while and it sends me a notification. It's like right time to stand up. I will usually listen to it and back, right. I'll just stretch my legs for 5 minutes and then come back’ (KL).  ‘Sleep 100% has got so much better I think because I know so much what not watching me sleep. But like there is something tracking my sleep. Like I'll always try and go to sleep earlier now. And I'll always sleep for longer now and like, if I know I'm not getting to bed on time, you know, when you get yourself all worked up because you're like, oh, it's 1:00 and I can't sleep. I think that's got worse because. In the same like kind of. I'm like, oh, I know something's tracking and I don't know. It's gonna give me a bad score in the morning, oh no’ (KL).  ‘If I'm just having one of them nights where I can't sleep, it almost has the kind of opposite effects and stresses you out more because you're like, I know, I know it's going to tell me off in the morning’ (KL). ‘If I have a normal night and can get to sleep and stay asleep, it's great. Like I know I'm getting a good score in the morning. I feel really excited about that. (KL).  ‘I've never got to the point where I've taken the watch off and just thought sod it, I'm not putting it on, but if I've forgotten to put my watch on for the night and then I'm struggling to sleep, it's almost kind of a relief because I'm well. At least I'm not going to get told off in the morning. (KL).  ‘I'll purposely not wear my watch because I know I won't be getting home at least an hour. Do you know what I mean? So I just don't want it to know about that. I will purposely take it off just so my graph still looks nice’ (KL). |
| Access to unlimited information | ‘I love my watch and knowing my steps but I intentionally keep it at that so it doesn’t get too much’ (JP).  ‘If I’m ill I’m bad at resting anyway, but I’ve got lupus which is, is just pretty nasty, and I’ve started taking my watch off if I’m in bed. I’m in bed and very much aware of that, and hating it! Yeah, it’s annoying and I’ve definitely found myself feeling guilty’ (JP).  ‘Too much information isn’t good in my eyes. You just don’t need all that’ (JP).  ‘In my mind I wouldn’t get obsessive about it, but I think it’d make me stressed. Yeah, stressed from the amount of numbers I got’. (JP).  ‘I keep it simple and I think that’s why I’m okay – not too much data so I’m not too affected’ (JP).  ‘If I’m having to have a slow day it feels like a reminder that I’m not doing great’ (JP).  ‘If I’m poorly, I know I wouldn’t be reminded I was doing bad without a smartwatch, whereas I’m wearing one and I feel bad about it telling me I’m not doing anything’ (MT).  ‘I do think I could get rid of maybe that guilt from a smartwatch, but I’d still know I wasn’t doing anything and the numbers attached to that so actually while sometimes seeing I’ve not done anything and my like fitness status is decreasing I would be aware of that now without a watch on’ (AH).  ‘Body freedom and just not having to think about things every single day is something I can’t really imagine’ (AH).  ‘Maybe after festivals or like weddings, where I’ve come back. I’ve drank quite a bit, it’s been emotionally taxing, it’s been a long day with bad sleep maybe it’s not helpful to have that information. As in, not pleasant, maybe it would be, what’s the saying, ignorance is bliss? Maybe it would be better to just not have access to that level of granularity in those circumstances’ (MH).  ‘Coming back to that ignorance is bliss, I definitely think wearing a smartwatch has had an impact on my relationship with alcohol, in that, I feel like I’m now so aware of the impact it has on my sleep and overall training readiness and I can’t but the cat back in the bag now with that. So I feel it’s probably created quite a bit of pressure to not drink there’ (MH).  ‘I don’t feel like it’s stolen anything from me, but on the other side of things I think I was so strict last year I did look back and tell myself it’s not that deep. Not like lingering resentment, but I’m having a debate over drinking and that was only from my watch.’ (MH).  ‘It’s equally brilliant and inconvenient information, I sometimes wish I didn’t know if that makes sense. Like I said though, the cats out the bag, I can’t unsee that now’ (MH).  ‘It’s just like awareness and you can’t un-know something’ (MH).  ‘I’m just pretty into it, I’m on it a lot and wear it all the time. I really enjoy looking at the data but I can sometimes find myself panicked on numbers like heart rate and steps when nothing has actually changed in the last year, I just have the information now’ (ZA).  ‘I was chatting to a friend and we said it was a bit like, do you remember doomscrolling in COVID? It was a bit like doomscrolling and fearmongering. It can feel a bit like that, like I’m, i’m almost actively looking at something to stress me out’ (ZA).  ‘I think over time from seeing the data and how it changes when I’m more active, it’s motivating me over time, but it’s probably come from the fact I feel pressured to stick to it. Like now I know, now I know how much movement actually impacts the calories burned and what my body needs it could be pressure’ (ZA).  ‘I think I just know a bit too much from it, but also that’s what I like and why I got it’ (ZA).  ‘I don’t know if now I’ve had one, if getting rid of my watch smartwatch would mean I’d just be trying to estimate myself and then panicking about my guesses. So I don’t know if now I’ve got one it would make me less anxious to take it off. You don’t know what you don’t know, but now I’ve worn one I know the impact of movement. I just don’t know if I’ve done 5,000 or 6,000 steps so I’d only start guessing I reckon. And yeah, I feel like I’d probably be more tired by thinking like that’ (ZA).  ‘I’m obsessed with my heart rate, probably to the detriment of my anxiety’ (MN).  ‘I feel like the watch is like the devil on my shoulder, or wrist. I feel like I’m telling myself about how I’m going to be more relaxed and then from having those numbers my brain is going but what, but what about if you miss a PB, what if you, yeah, it’s messy’ (LP). It kind of makes you not a second guess, but kind of makes you forget about the bigger picture. Oh, it doesn't matter that I slept 6:50 and I'm really happy about that. Instead it kind of makes you zone in on the small things like what happened at 1:20am?’ (KL). |
| Perceived success comes with compromise | ‘I feel like I’ve not recognised the good effects it has on me and my steps might come out of guilt which is messy to unpack’ (JP).  ‘It’s not great, but I don’t know. I feel like my smart watch makes me better because it, it makes me push myself so… I don’t know. It’s bad and good. I’d say mainly bad but I do like it’ (MT).  ‘On paper it might have made me physically healthier, but it made me more obsessive with numbers too’ (AH).  You’re kind to yourself, you’re healthier, incredible! And I agree with that but I’m aware at the moment I’m very dependent on it’ (AH).  ‘While there could be all of these negative impacts, wearing a smartwatch makes me healthier’ (AH).  ‘I know a lot of people that just take them off for months at a time because they can’t like deal with the reminders and stuff, and then they put it back on when they feel good, it’s like a viscous cycle… you’re not going to get any results if you just keep taking it off and putting it back on, like they need to pick a middle ground’ (HG).  ‘I really like that I get notifications, and I like and use all the fitness stuff… it’s good, but it’s a bit toxic. It, I, t think it’s a bit like in a relationship, a toxic relationship. You’re in it but it might not be best you. That’s what my girlfriend jokes at me’ (ZA).  ‘Wearing a watch does make me a fitter person. It makes me more active, which is why I got the watch, what I set out to do. And I think it makes me more competitive with those numbers, so I hate it and it can be a little unhealthy but the watch is having the effect I wanted. So yeah, I still wear my watch and do like it’ (ZA).  ‘I think it makes me quite anxious having the data, but it’s the anxiety that brings, is what makes me do it, so it’s kind of like a mean teacher that I don’t like, but then I’ve opened my results and think you know what, they knew what they were doing’ (ZA).  ‘I can’t say it’s not toxic because I’m putting a lot of trust in this in this watch and the questions you’ve asked have made me say out loud that I’m putting the watch data over my own mind, but it’s really good for my goals so I think I’ll just shrug it off for a bit’ (ZA).  ‘It’s just so good for me physically but is probably really bad for me mentally too’ (ZA).  ‘Maybe less motivated, erm, I think I’d probably do a bit less, like I do think it wouldn’t tell me off being being quite sluggish which, again, could be good, but it’s not what I need right now. I’d probably be a bit more relaxed but also I wouldn’t be as fit’ (ZA).  ‘I don’t think there are any like positives from day-to-day… I mean surely there is. There must be if I wear it, right? But emotional wise I’m struggling to think of them’ (MN). |
